# The Impact of Non-coding G-quadruplex Variants on Human Traits and Disease Susceptibility

**DOI:** 10.64898/2026.05.29.26354456

**Authors:** Rahul Sharma, Fengyuan Hu, Xiaoyin Li, Rafael Campos, Kousik Kundu, Santosh Atanur, Marcin Karpinski, Sebastian Wasilewski, Stewart MacArthur, Dimitrios Vitsios, Ryan S. Dhindsa, Ilias Georgakopoulos-Soares, Oliver S. Burren, Slavé Petrovski, Anthony M. Mustoe, Quanli Wang, Dominik Glodzik, Xueqing Zoe Zou

## Abstract

Non-coding variants are important contributors to human traits and diseases but linking them to molecular mechanisms and phenotypes at scale remains challenging. G-quadruplexes (G4s) are four-stranded structures formed by guanine-rich sequences and have emerged as key functional elements within the non-coding genome. G4s are enriched in regulatory regions and can modulate gene expression at both the DNA and RNA levels, influencing transcription, replication, and RNA processing, positioning them as key mediators linking non-coding variation to complex biological traits. Here, we profile putative G4s across five regulatory regions in 459,449 UK Biobank genomes and perform phenome-wide association analyses spanning 2,941 plasma protein abundances, 13,321 binary traits, and 1,682 quantitative traits. We show that putative G4-modifying variants are depleted under purifying selection despite elevated local mutability and drive large, bidirectional associations with plasma proteins and clinical traits, including associations not captured by coding variants. Using a mechanism-aware collapsing strategy that groups rare non-coding variants by their predicted impact on G4 stability, we achieved stronger gene-level signals than those obtained with standard rare-variant collapsing approaches. Integrating non-coding and protein-truncating variants (PTVs) increases discovery power, revealing 843 significant associations missed by the PTV-only model. Replication in the Alliance for Genomic Discovery cohort demonstrates cross-cohort robustness. Our study suggests G4s as widespread mediators of non-coding regulation and provides a framework for mechanism-informed target discovery and prioritization across the non-coding genome.

## Introduction

The non-coding genome plays a central role in gene regulation [1], and non-coding variants contribute to human traits and disease susceptibility [2–4]. For example, a variant in 3*′* untranslated region (3*′*UTR) of *APOA5* has been associated with reduced APOA5 protein levels, elevating triglycerides and hypertriglyceridemia risk [5]. Recent genome-wide association studies have shown that non-coding variants can influence circulating protein levels as strongly as coding variants [6, 7]. These functional non-coding variants are found to be enriched in known regulatory regions such as UTRs, promoters, and enhancers [6–8]. Yet, for most non-coding associations, the regulatory mechanism remains unclear, limiting biological interpretation and translational use. One underexplored axis of regulation is non-canonical nucleic acid secondary structure. More than 13% of the human genome can adopt non-B DNA structures (i.e., structures other than the canonical double helix) [9–14]. Among these, G-quadruplexes (G4s), four-stranded structures formed by stacked guanine tetrads and stabilized by cations [11, 15], are widespread in regulatory regions [16, 17], have been visualized in human cells [18], and mapped genome-wide [12, 13, 19]. Importantly, G4s can exert regulatory effects at both the DNA and RNA levels by influencing transcription [15, 20–23] and replication [24, 25] when folded in DNA, and by modulating translation and RNA stability when formed in transcribed sequences [26].

Noncoding variants within regulatory region G4s can thus exert their effects through modulation of G4 structures, affecting gene expression and potentially causing phenotypic variation [16, 20, 27–31]. For example, alteration of a DNA G4 structure by a single-nucleotide variant in the *c-MYC* promoter led to a threefold increase in c-MYC expression [31]. Alternatively, RNA G4-formative and G4-disruptive variants in the 5*′*UTR of *MSH6* and *EPN3* reduced their protein expression [32], underscoring versatile and context-dependent G4 regulation. G4 dysregulation has been linked to cancer [33] and other diseases including neurodegenerative diseases [34]. G4s are also attractive therapeutic targets [35–39]: several G4-targeting compounds exemplify this potential including BRACO-19 [40], RHPS4 [41], and CX-5461 [42]. Furthermore, diverse precision therapeutic approaches, including small-molecule G4 modulators [43], inhibition of G4-associated proteins [44], antisense oligonucleotide approaches [45, 46], and CRISPR–Cas genome editing [47], demonstrate both the feasibility and the precision of G4-directed therapies.

While over a half-million G4s have been computationally predicted or experimentally identified in human genome [13], only a subset is likely to be functional, such that their disruption has phenotypic effects. Distinguishing functional from non-functional G4s remains a major challenge. Phenome-wide association studies (PheWAS) offer a scalable framework to link G4-modifying variants with phenotypic outcomes, thereby enabling identification of functional G4s. However, most prior PheWAS have focused on coding variants [48–51], with only recent work has begun interrogating non-coding regions [6, 7, 52, 53], including studies of RNA-G4 variants in breast cancer [32] and DNA-G4 variants linked to metabolic diseases [54]. A key methodological gap constraining functional exploration of the non-coding genome is that the gene-based collapsing analyses are particularly suited for coding variants (aggregate rare variants with largely loss-of-function effects e.g., PTVs [48, 49, 53]), whereas non-coding variants often exert bidirectional effects [7], leading to signal cancellation and reduced statistical power under naive aggregation. Here, we propose a mechanism-aware collapsing framework, grouping variants by their predicted impact on G4 structure, that can preserve directional coherence and enhance interpretability, and systematically evaluate the framework at population-scale.

In this study, we conducted a systematic, population-scale PheWAS in nearly half a million UK Biobank (UKB) genomes with a mechanism-aware collapsing approach that groups rare non-coding variants by their predicted impact on G4 stability. We first mapped putative G4s (PQs) across five key non-coding regulatory elements (hereafter referred to as *regulatory regions*), including promoters (200 base pair (bp) upstream of transcription start sites), splice-proximal introns, CTCF binding sites (defined as 200 bp windows centered on each CTCF binding site), 5*′*UTRs, and 3*′*UTRs and classified variants as putative G4-disruptive (removing a G4), G4-formative (creating a G4), or G4-modulatory (changing G-tract or loop length/count while retaining the G4) (Fig. 1a). We then performed variant-level association tests of PQ variants in *regulatory regions*, together with non-G4 variants in promoters and CTCF binding sites and conducted gene-level collapsing analysis in *regulatory regions* across plasma protein abundances (2,941 proteins), 13,321 binary traits, and 1,682 quantitative traits. At the variant level, PQ-modifying variants drive large, bidirectional associations with proteins and traits, including signals not captured by coding variants. At the gene level, collapsing by predicted G4 mechanism yields stronger and more interpretable associations than standard non-coding burden tests. Integrating non-coding PQ variants with PTVs uncovers associations missed by either class alone. Finally, we test the generalizability of our findings in an independent cohort from the Alliance for Genomic Discovery. Collectively, these results provide new mechanistic insights into functional consequences of G4 variants and support a mechanism-informed framework for target discovery and prioritization in non-coding regions.

**Fig. 1:**
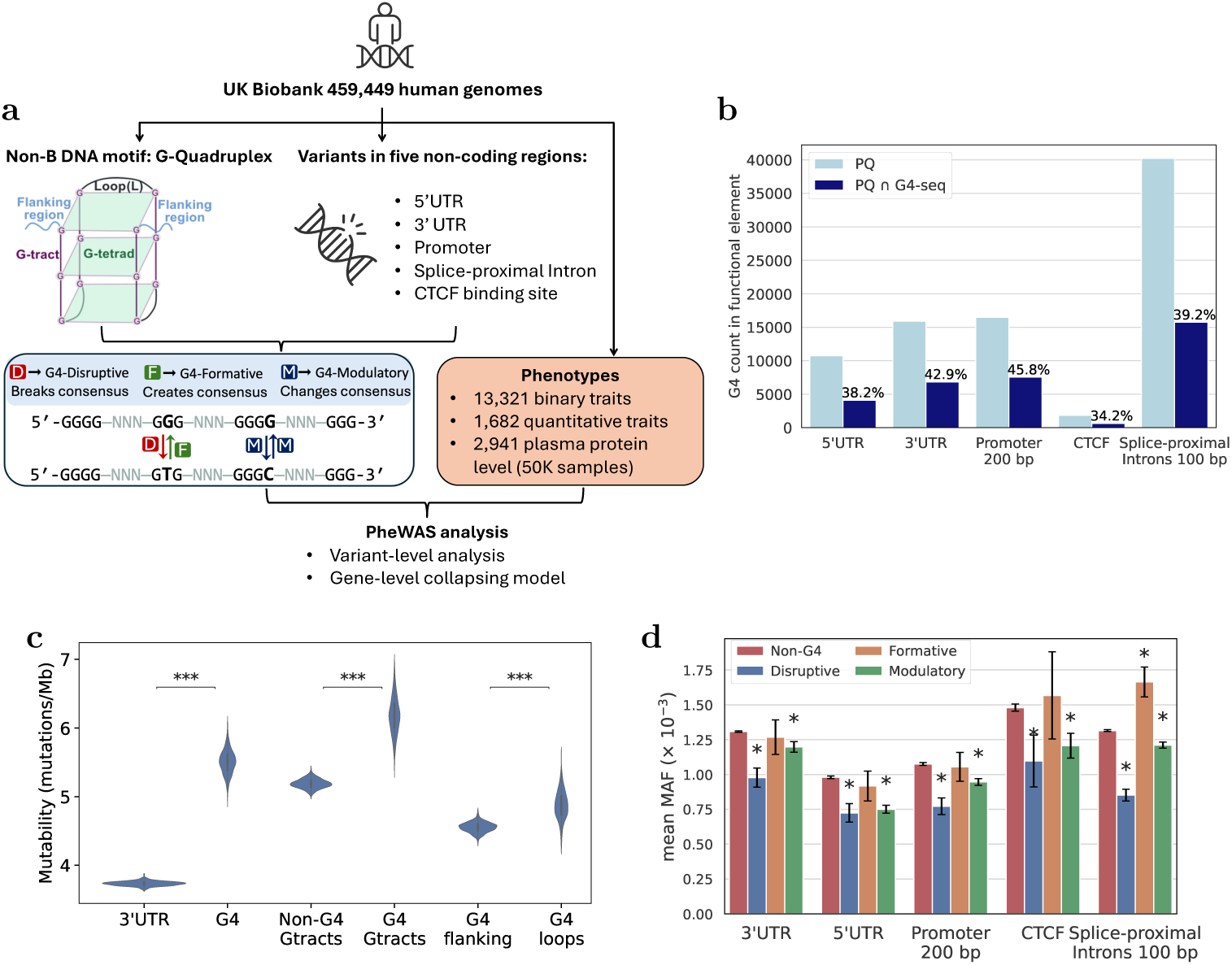
Study overview and G-quadruplex (G4) features. **(a)** Analysis framework for phenome-wide association studies (PheWAS) of putative G4 (PQ) variants in the UK Biobank 459,449 WGS. PQ motifs were mapped within five non-coding regulatory regions: 5*′*UTR, 3*′*UTR, promoter, splice-proximal intron, and CTCF binding site. PQ variants were classified as disruptive, formative, or modulatory (Methods) and were tested for association with 2,941 plasma protein abundances (measured in ∼50,000 samples), 13,321 binary traits, and 1,682 quantitative traits. Association analyses were conducted at both variant- and gene-level. **(b)** Bar plot showing counts of putative G4s (PQs) and G4s supported by high-throughput G4-seq experiments [13], both matching the {G_3+_L_1-12_}_3+_G_3+_ motif (G denotes Guanine and L is loop of 1-12 nucleotides of any base). The overlap between PQs and G4-seq G4s is indicated above the dark blue bars. **(c)** Mutability comparison of different regions, including all 3*′*UTR regions vs 3*′*UTR G4 regions, G-tract regions in G4 vs in non-G4, G4 loops vs G4 flanking sequence. Distributions were obtained from 1000 subsampling iterations and significance (two-sided t-test) is indicated by asterisks where (∗ ∗ ∗) denotes *p <* 10*^−^*^150^. Results for other *regulatory regions* are in SI Fig. 2a-d. Trimer frequency contexts in G4 loops and flanking regions in SI Fig. 2e-g. **(d)** Distribution of mean MAF (minor allele frequency) of PQ variants compared to non-PQ variants stratified by functional class. Error bars denote the standard error of the mean. The ∗ indicates a statistically significant difference (Welch’s t-test, *p <* 0.05) in mean MAF between PQ and non-PQ variants within the same non-coding regulatory region. Icons in this figure were adapted from Flaticon.com.

## Results

### Widespread G-quadruplexes are more mutable but subject to strong purifying selection

We identified 676,611 PQs genome-wide using the quadparser algorithm [11] (Methods), with 101,429 (∼15%) falling within the examined *regulatory regions* (SI Table 1). Approximately 41% of PQs in *regulatory regions* were supported by G4-seq experiments [13] (Fig. 1b). Notably, 86.7% of genes contain at least one PQ motif within their *regulatory regions* (Table 1 and SI Fig. 1a-e), with at least one PQ motif present in 62.9% of 5*′*UTR, 42.1% of 3*′*UTR, 61.9% of promoters (200 bp), 73.9% of splice-proximal introns and 21.9% of CTCF binding sites. Most PQs are short (median length 35 bp), contain six or fewer G-tracts (SI Fig. 1f), typically (∼87%) with three or four Gs per tract (SI Fig. 1g), and have loops shorter than 8 bp in ∼77% of PQs (SI Fig. 1h).

**Table 1:** Counts of genes with putative G4s (PQs) in their non-coding regulatory regions and PQ variants in those regions. Percentages are relative to the total number of genes associated with each specific regulatory region. Overlaps among genes harbouring various non-coding regulatory regions, genes with PQs in those regulatory regions, and genes with PQ variants in those regulatory regions are shown in UpSet plots in SI Fig. 1d-e and SI Fig. 3f-g.

|  | PQ variants<br>category | 5’UTRs | 3’UTRs | Promoters<br>(200 bp) | Splice-proximal<br>Introns |
| --- | --- | --- | --- | --- | --- |
| **Genes** |  | 18306 | 18310 | 18736 | 17754 |
| **Genes**<br>**with PQ** |  | 11515<br>(62.9%) | 7714<br>(42.1%) | 11605<br>(61.9%) | 13128<br>(73.9%) |
|  | G4 disruptive | 5899<br>(32.2%) | 5992<br>(32.7%) | 8061<br>(43.1%) | 10023<br>(56.5%) |
|  | G4 formative | 5454<br>(29.8%) | 4382<br>(23.9%) | 6305<br>(33.7%) | 8067<br>(45.4%) |
|  | formative & disruptive | 3391<br>(18.5%) | 3740<br>(20.4%) | 4557<br>(24.3%) | 6765<br>(38.1%) |
| **Genes**<br>**without PQ** |  | 6791<br>(37.1%) | 10596<br>(57.9%) | 7131<br>(38.1%) | 4626<br>(26.1%) |
|  | G4 formative | 1375<br>(7.5%) | 1966<br>(10.7%) | 2140<br>(11.4%) | 1213<br>(6.8%) |

Variant distributions across the allele frequency spectrum reflect the interplay between local mutability, which determines the input of new variants, and selection, which is indicative of functional importance. To disentangle mutational propensity from selection in PQ regions, we first computed local mutability, defined as the average burden of rare variants (minor allele frequency, MAF *<* 0.1%) per base pair (Methods). We observed elevated mutability in PQs relative to their underlying regulatory regions; for instance, 5.51 mutations were observed per megabase (Mb) in 3*′*UTR PQ regions versus 3.73 mutations/Mb in 3*′*UTR (two-sided t-test, *p <* 10*^−^*^150^). Similar patterns were observed in G4 G-tracts versus non-G4 G-tracts and G4 loops versus G4 flanking regions (Fig. 1c) across *regulatory regions* (SI Fig. 2) and in the independent dbSNP dataset [20]. The high mutability of G4 regions can be attributed to single-stranded DNA conformations adopted within G4 structures [20] or to co-occurring R-loops [55].

We then categorised PQ variants into Disruptive, Formative, and Modulatory based on whether they disrupt, create, or modulate (i.e., alter G-tract or loop length/-count without disrupting) the PQ consensus motif and thereby providing an estimate of their effect on PQ stability (see Methods). Across *regulatory regions*, G4-modifying variants were approximately 80% Modulatory, 14% Disruptive, and 6% Formative (SI Fig. 3a; SI Tables 2). Approximately 79% of all protein-coding genes contain G4-disruptive variants in their *regulatory regions* (Table 1 and SI Fig. 3(b-g)), while over 51% of genes without G4s have G4-formative variants. Notably, despite the high mutability of G4 regions, G4-modifying variants are relatively rare (*>*98% with MAF *<* 0.1%) (SI Fig. 4a). In particular, G4-disruptive and G4-modulatory variants have significantly lower mean MAF than non-G4 variants in the same regions (Welch’s t-test, *p <* 0.05), including within *regulatory regions* of intolerant genes which already show depletion of non-coding variation (Fig. 1d, SI Fig. 4b), implying their efficient removal by purifying selection. These frequency skews replicate in an independent cohort (SI Fig. 4c-d) and align with prior reports [16, 32, 56].

High mutability coupled with low allele frequencies indicates purifying selection, positioning putative G4-modifying variants as compelling regulation mediators. This prevalence and constraint motivate us to test whether variants that form, disrupt, or modulate G4s drive phenotypic associations.

### Non-coding G4 variation drives large-effect, bidirectional, coding-independent associations

To test whether PQ-modifying variants have phenotypic consequences, we conducted association analyses for putative G4-modifying variants located within *regulatory regions* and non-G4 variants within 200 bp upstream of promoter regions and within 200 bp windows centered on CTCF binding sites (Methods).

We first assessed the association of variants with 2,941 plasma protein levels (SI Table 3) in about 50k individuals. Using a significance threshold of *p* = 10*^−^*^8^ (false discovery rate (FDR) = 0.02%), we identified 1,175 *cis*-protein quantitative trait locus (pQTL) associations (74 rare (MAF *<* 0.1%) and 1,101 common) and 15,073 *trans*-pQTL associations (408 rare and 14,665 common) attributable to 6,380 G4 variants in *regulatory regions* (Fig. 2a-b, SI Fig. 5, SI Tables 4-5). As expected, rare variants exhibit substantially larger effect sizes (*β*) compared to common variants (|*β*|_mean_: 1.33 ± 0.58 versus 0.21 ± 0.22; SI Fig. 6a). Consistent with prior work [7], non-coding *cis*-pQTLs show a more balanced direction of increasing and decreasing protein levels (46.9% increasing among common; 51.3% increasing among rare) compared with protein-truncating variants (PTVs) which predominantly decrease protein levels (16.5% increasing among common and 3.1% increasing among rare [49]) (SI Fig. 6b-c, SI Table 6). *Trans*-pQTL associations were balanced for both coding and non-coding variants (SI Fig. 6b-c, SI Table 6).

**Fig. 2:**
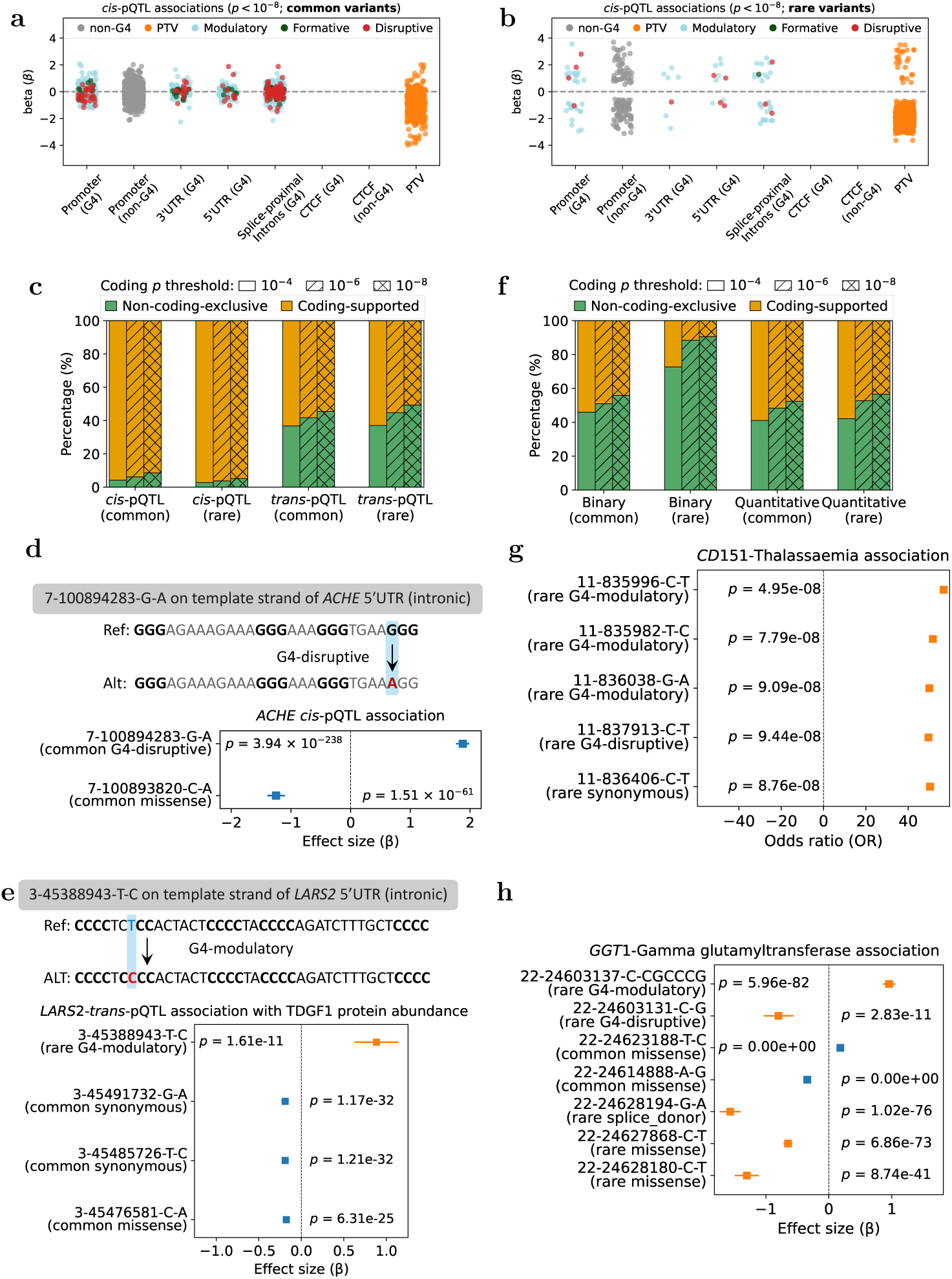
Non-coding variant-level association analyses summary. **(a-b)** Effect size (*β*) distribution of *cis*-pQTLs for **(a)** common and **(b)** rare (MAF *<* 0.1%) variants in *regulatory regions*, alongside protein-truncating variant (PTV) associations [48, 49, 53]. Colored points denote different classes of variants (non-G4 non-coding, G4-modulatory non-coding, G4-disruptive non-coding, G4-formative non-coding, and protein-truncating coding variants). **(c)** Proportion of non-coding-exclusive signals (green) or coding-supported signals (orange) for significant noncoding *cis* and *trans* pQTLs separated by common and rare variants. Bars are stacked by category: green indicates non-coding-exclusive associations; orange indicates coding-supported associations. Hatch patterns denote the coding significance thresholds (*p <* 10*^−^*^4^, *p <* 10*^−^*^6^, *p <* 10*^−^*^8^). **(d-e)** Forest plots comparing effect sizes across multiple coding (synonymous, missense) and non-coding (G4-modulatory, G4-disruptive) variants for *cis*-pQTL associations in *ACHE* **(d)** and *trans*-pQTL associations of *LARS2* variants with TDGF1 protein abundance **(e)**. **(f)** Proportion of non-coding-exclusive signals (green) or coding-supported signals (orange) for significant noncoding binary and quantitative traits separated by common and rare variants. **(g-h)** Forest plots of effect sizes for coding and non-coding variants: **(g)** *CD151* variants associated with Thalas-saemia and **(h)** *GGT1* variants associated with gamma glutamyltransferase. In forest plots, rare and common variants are highlighted in orange and blue, respectively. G4 sequence context for these variants are listed in SI Table 9.

Most non-coding *cis*-pQTL signals (common 91.3%; rare 94.8%) are also observed for coding variation in the same gene (referred to as coding-supported). In contrast, coding support was lower in *trans*-pQTL signals (common 54.6%; rare 50.7%) (Fig. 2c), likely reflecting stronger and more direct regulatory mechanisms in *cis*-than in *trans*-pQTL associations. Among non-coding-exclusive pQTLs, G4 variants accounted for 89 (37.1%) *cis*- and 5,983 (36.2%) *trans*-pQTLs. For example, a rare G4-modulatory promoter variant (19-676317-GCCGCGCCCCGCC-G) in *FSTL3* was identified as a *cis*-pQTL (*p* = 8.97×10*^−^*^19^; *β* = −1.27) at variant-level, yet no coding *cis*-pQTLs were detected in *FSTL3* (SI Fig. 7). Similar proportions of non-coding-exclusive and coding-supported pQTL associations were observed across *regulatory regions* (SI Figs. 8-9).

Notably, some non-coding G4 variants exert effects on protein levels opposite to coding variants within the same gene. For instance, in *ACHE*, a 5*′*UTR (intronic) G4-disruptive variant (7-100894283-G-A) on the template strand is associated with increased ACHE protein level (*p* = 3.94 × 10*^−^*^238^; *β* = 1.88) whereas a missense (7-100893820-C-A: (*p* = 1.51 × 10*^−^*^61^; *β* = −1.25) variant exhibits opposing effects (Fig. 2d). In trans-pQTLs, a rare G4-modulatory variant (3-45388943-T-C) in the 5’UTR (intronic) of *LARS2* is associated with increased TDGF1 protein abundance (*p* = 1.61 × 10*^−^*^11^; *β* = 0.88). *LARS2* also contains two synonymous and one missense variants showing the same *trans*-pQTL signal but with decreased effects (Fig. 2e). These examples support that G4-mediated regulatory mechanisms may operate independent of coding variants and, in some contexts, can produce opposite functional consequences.

We next examined these molecular effects at the phenotypic level. Across 13,321 binary and 1,682 quantitative traits (SI Table 3), G4-modifying variants were involved in 2,103 significant binary and 19,457 quantitative associations (SI Fig. 10-11, SI Tables 7-8). For common variants, 44.2% (2,512/5,686) of binary and 47.5% (23,678/49,714) of quantitative associations were coding-supported; for rare variants, 9.5% (9/95) of binary and 43.5% (218/501) of quantitative associations were coding-supported (Fig. 2f; SI Fig. 8), with analogous proportions for G4 variants (SI Fig. 9). Among non-coding-exclusive associations, G4 variants accounted for 1,103 (33.8%) binary and 8,984 (34.1%) quantitative associations.

SI Fig. 12a highlights loci with the strongest binary associations across major disease domains. For example, a rare 5*′*UTR (intronic) G4-modulatory variant (14-24310089-A-AGGGAGGGGCCCC) on the non-template strand of *LTB4R2* is associated with primary muscle disorders (*p* = 5.1 × 10*^−^*^9^; OR = 2, 776.46). *LTB4R2* encodes a G protein-coupled receptor for the lipid mediator leukotriene B4 [57] and is involved in recruiting and activating immune cells during inflammatory responses [58]. Because dysregulated inflammation is a common feature in many primary muscle disorders, including muscular dystrophies, this association is consistent with a role of inflammatory regulation in muscle pathology. Notably, no coding variants in *LTB4R2* showed a strong association with this phenotype. Another interesting example is the association of three rare G4-modulatory and one rare G4-disruptive variants in the splice-proximal intron of the *CD151* gene with Thalassaemia alongside a rare synonymous coding variant [48] (Fig. 2g, SI Table 9). Notably, *CD151* -deficiency requires increased erythropoietin in *β*-thalassemia [59], suggesting its potential role in regulating treatment response. Together, these findings suggest that non-coding variants, such as G4 variants, may contribute to disease risk and warrant further functional investigation to disentangle non-coding regulation from coding contributions.

SI Fig. 12b shows quantitative associations, with the strongest variant-trait pairs exemplifying both positive and negative directions of effect. For example, a rare G4-modulatory variant located in the 5*′*UTR (intronic) on the non-template strand of *ASGR1* (encodes the major subunit of the Ashwell-Morell receptor that removes circulating glycoproteins [60]) is associated with increased alkaline phosphatase levels (17-7179137-T-C: *p* = 5.38 × 10*^−^*^9^; *β* = 1.31; SI Fig. 13). Previous PheWAS identified coding and intronic *ASGR1* variants associated with elevated alkaline phosphatase [48, 61]. Another example is the association between promoter G4 variants in *GGT1* and gamma-glutamyltransferase: a rare G4-modulatory variant (extending G4) is associated with increased gamma-glutamyltransferase level (22-24603137-C-CGCCCG: *p* = 5.96 × 10*^−^*^82^ and *β* = 0.92), whereas a nearby G4-disruptive variant decreased levels (22-24603131-C-G: *p* = 2.82 × 10*^−^*^11^; *β* = −0.80), alongside four missense and one splice-donor coding variants in *GGT1* (*p <* 10*^−^*^40^) (Fig. 2h, SI Table 9).

Finally, to assess whether significantly associated non-coding variants were tagging previously known signals, we evaluated their linkage disequilibrium (LD) with nearby GWAS variants reported in the NHGRI-EBI GWAS Catalog (see Methods). Among significantly associated rare non-coding variants, 52 of 1,440 (3.6%) were in LD (R^2^ ≥ 0.2) with previously reported GWAS variants located within 500 kb on either side and associated with the same phenotype. As expected, the corresponding proportion was substantially higher for common non-coding variants, with 9,583 of 19,420 (49.3%) meeting the same criterion (SI Tables 10-11).

### Functional non-coding G4s are enriched among G4-seq-supported smaller G4s and show no strand bias

Variant-level PheWAS showed that non-coding G4-modifying variants are broadly associated with *cis*- and *trans*-acting molecular phenotypes and diverse traits, frequently with large and bidirectional effects. We next defined “functional” G4s as those carrying PQ-modifying variants significantly associated with any UK Biobank phenotype. Fig. 3a shows the distribution of these functional G4s across *regulatory regions*. Of the 101,429 G4s identified in *regulatory regions*, 67,936 (68.0%) harboured G4-modifying variants observed in at least five samples and were included in the association analysis. Of these 67,936 G4s, 6,640 (9.8%) were functional (Fig. 3b). Notably, 1,771 of 12,506 (14.2%) G4-seq-supported G4s with shorter loops (≤ 7 nucleotides) were functional, representing a significant enrichment relative to other G4s (OR = 1.71 and *p* = 1.14 × 10*^−^*^68^, two-sided Fisher’s exact test). In addition to existing functional G4s, we identified 495 unique G4-formative variants that create new functional G4s, including 100 in promoters (200 bp), 68 in 5*′*UTR, 100 in 3*′*UTR, 13 in CTCF binding sites and 242 in splice-proximal introns.

**Fig. 3:**
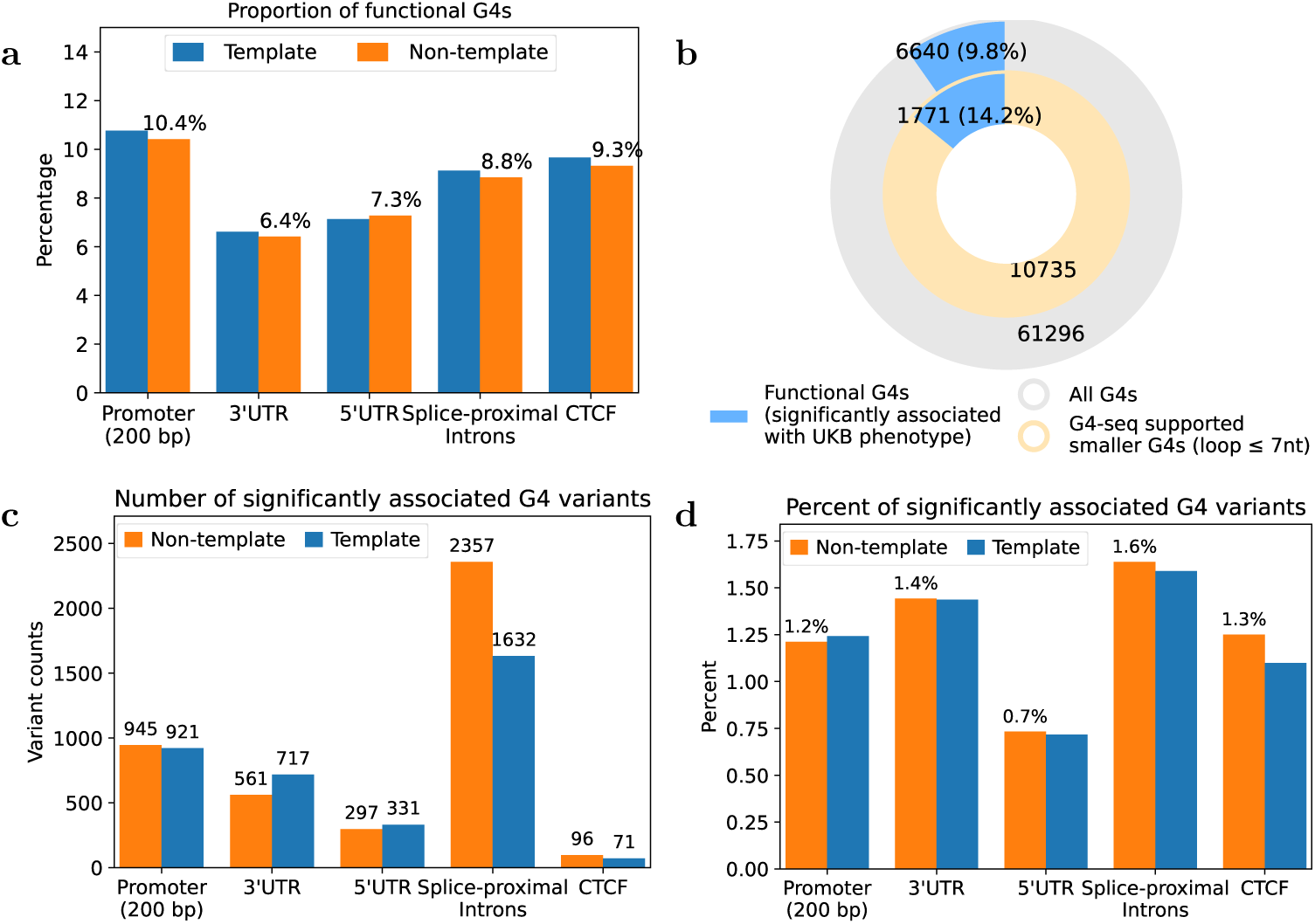
Distribution of functional G4s and associated G4 variants across template and non-template strands in *regulatory regions*. **(a)** Percentage of functional G4s across *regulatory regions*, stratified by their distribution on the template and non-template strands. Functional G4s are defined as PQs containing at least one PQ-modifying variant significantly associated with any phenotype in the UK Biobank dataset analyzed in this study. **(b)** Proportion of functional G4s among all PQs (outer ring) and among G4-seq supported PQs with shorter loops (≤ 7 nucleotides), highlighting enrichment within the smaller G4 subset. **(c)** Count of significantly associated G4 variants on the template versus non-template strands across *regulatory regions*. **(d)** Percentage of significantly associated G4 variants on the template versus non-template strands across *regulatory regions*. Note: All analyses consider only PQ variants with at least five carriers or PQs that contain such variants.

We next sought to understand the potential mechanisms through which these PQs may act. PQs located within promoters and CTCF binding site likely function at the DNA level, and significantly associated promoter and CTCF variants are balanced between the template and non-template strands, suggesting a lack of strand specificity (Fig. 3c-d). By comparison, PQs located in UTRs or splice-proximal introns may function at either the DNA level, or, when located on the non-template strand, at the RNA level. Again, the distribution of significantly associated variants is balanced between the template and non-template strands, with no evident bias towards an RNA-based mechanism (Fig. 3c-d). However, we note that, since PQ sequences were identified from genomic DNA, our analysis is not powered to detect PQs and their variants in spliced transcripts that may function at the RNA level.

### *G4-Mechanistic non-coding* collapsing reveals larger effect associations than conventional rare-variant collapsing

*Regulatory regions* of a gene often contain multiple G4-modifying variants (SI Table 12 and SI Fig. 14). For example, UTRs contain on average 9.8 G4-disruptive, 4.2 G4-formative, and 29.4 G4-modulatory variants per gene. Mirroring the logic of gene-based collapsing models of coding variants [48], aggregating multiple non-coding variants within the same regulatory regions may reveal gene-level associations that are undetectable at the single-variant level. We therefore conducted gene-level collapsing analyses (Fig. 4a) by grouping rare (MAF *<* 0.1%) qualifying variants within promoter (200 bp), promoter (1 kb), splice-proximal intron, UTR, and combined UTR–promoter (200 bp) and UTR-promoter (1 kb) regions. Combining UTR and promoter regions reflects their integrated role in cis-regulation, jointly modulating transcription, mRNA stability, and translation, and therefore, attempting to model the total coherent regulatory burden that affects gene dosage. For each of the six regions, we applied five distinct collapsing strategies based on distinct qualifying-variant definitions: two *Baseline non-coding* strategies collapsing (a) all rare variants and (b) rare variants with CADD score ≥ 5, and three *G4-Mechanistic non-coding* strategies collapsing (c) G4-modifying rare variants, (d) G4-disruptive rare variants, and (e) G4-formative rare variants (SI Table 13, see Methods).

**Fig. 4:**
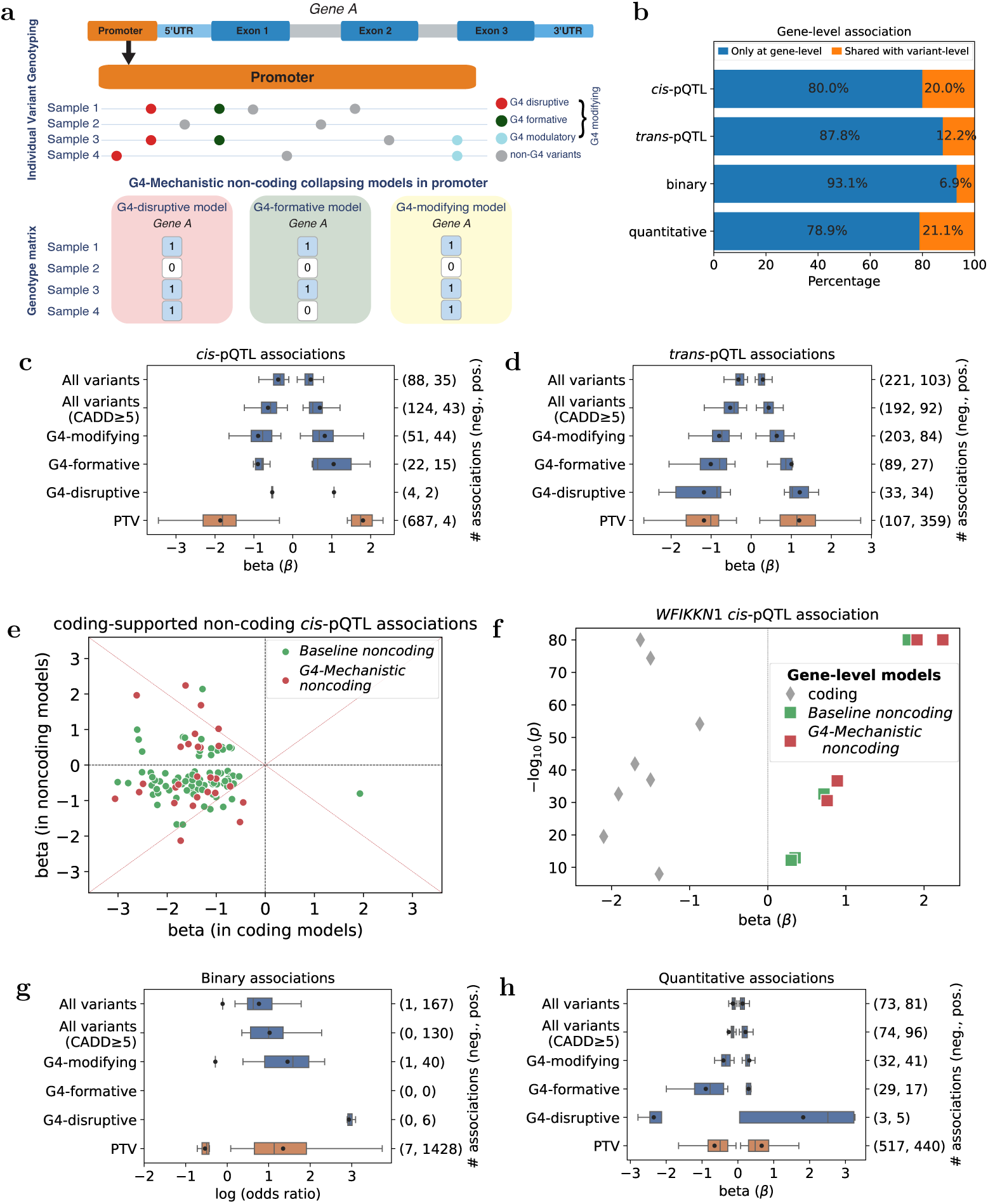
Non-coding gene-level collapsing model association summary. **(a)** Schematic illustration of the collapsing framework for three *G4-Mechanistic non-coding* models in regulatory regions, shown here for promoters. Top: promoter variants across four samples are classified by their predicted functional relationship to G-quadruplex (G4) structures: G4-disruptive (red), G4-formative (dark green), G4-modulatory (light blue), and non-G4 (grey). G4-disruptive, G4-formative, and G4-modulatory variants are collectively referred to as G4-modifying variants. Bottom: these variants are collapsed into binary genotype matrices for each *G4-Mechanistic non-coding* model. In the G4-disruptive model, a sample is scored 1 if it carries at least one G4-disruptive variant in the gene’s promoter. In the G4-formative model, a sample is scored 1 if it carries at least one G4-formative variant. In the G4-modifying model, a sample is scored 1 if it carries any G4-modifying variant. This study additionally implements two *Baseline non-coding* models (which collapse all rare variants and all rare variants with CADD ≥ 5) (SI Table 13). **(b)** Proportion of associations uniquely identified by the collapsing model (no individual variants included in that model achieved even marginal significance (*p <* 10*^−^*^4^) in the variant-level analysis) and also identified at variant-level analysis. **(c-d)** Distribution of effect sizes for significant *cis*-**(c)** and *trans*-pQTL **(d)** associations for non-coding collapsing models across various *regulatory regions* (SI Table 13) alongside coding protein-truncating variant (PTV) collapsing model (Methods; [49]). Positive and negative effects are shown separately (right-hand bar counts report the number of significant associations as (negative, positive)). **(e)** Effect sizes for coding-supported *cis*-pQTL associations (*p <* 10*^−^*^8^) observed in both coding and non-coding models, highlighting opposite effect associations between the two models. **(f)** Comparison of *WFIKKN1 cis*-pQTL signals for both coding (Methods; [49]) and non-coding (SI Table 13) gene-level association analyses. **(g-h)** Effect sizes for significant binary and quantitative associations for non-coding collapsing models of various *regulatory regions* and PTV collapsing model [48, 53].

Indeed, collapsing models increased power to detect signals: a substantial fraction of pQTL associations (∼86%) were exclusive to the non-coding collapsing models (Fig. 4b), with no single non-coding variant in the corresponding *regulatory regions* reaching marginal significance (*p <* 10*^−^*^4^). For example, a *trans* association between *IFRD2* gene and ACY1 protein abundance was detectable only in the *IFRD2* promoter (1 kb) G4-modifying model (*p* = 6.11×10*^−^*^17^; *β* = −0.97). In several loci with individually significant variants, collapsing further strengthened signals. For example, three G4-modifying variants in the *SPINK2* promoter were individually associated with increased SPINK2 protein abundance: 4-56821765-G-A (*p* = 2.22 × 10*^−^*^21^; *β* = 1.40), 4-56821765-G-GGGAAGGGGCGGA (*p* = 2.03 × 10*^−^*^14^; *β* = 1.22), and 4-56821735-G-T (*p* = 6.73 × 10*^−^*^9^; *β* = 1.19). The promoter (200 bp) G4-modifying collapsing model revealed even stronger association (*p* = 1.40 × 10*^−^*^39^; *β* = 0.88; SI Fig. 15). These results underscore the substantial statistical power gained through G4-informed collapsing models.

At genome-wide significance (*p <* 10*^−^*^8^, FDR = 1.46%), we identified 579 pQTL associations across all models, comprising 135 *cis*- and 444 *trans*-pQTL associations (SI Table 14, SI Fig. 16-17). Similar to variant-level pQTL associations, gene-level associations are also bidirectional (Fig. 4c-d). Notably, absolute effect sizes increase systematically from *Baseline* to *G4-Mechanistic* models, with the largest effects in the G4-formative model (1.16 ± 0.50), followed by G4-disruptive (1.00 ± 0.44), G4-modifying (0.78 ± 0.38), all-variant (CADD ≥ 5) (0.55 ± 0.30), and all variant models (0.33 ± 0.19) (Fig. 4c-d and SI Table 15). This pattern indicates that G4-informed collapsing models enrich for functionally coherent variants and reduce effect cancellation.

Comparing non-coding collapsing models with coding collapsing models [48, 49, 53] (Methods), we found that 124 (21.4%) non-coding pQTL associations were also observed in coding models (coding-supported), including 109 *cis* and 15 *trans* associations (SI Fig. 18a). Among these 124 associations, 26 showed greater significance, 18 showed larger effect and 9 showed both greater significance and larger effect in the non-coding models than coding models. For example, *RETN* showed a stronger *cis* association in the promoter (200 bp) G4 model (*p* = 1.77 × 10*^−^*^17^; *β* = −2.13) than in the coding model (ptvraredmg: *p* = 5.82 × 10*^−^*^12^; *β* = −1.72; Methods) [49]. Notably, ∼26% coding-supported *cis* significant associations had opposite effect directions between coding and non-coding models (Fig. 4e). For instance, in *WFIKKN1*, the UTR G4-modifying model was associated with increased protein levels (*p* = 1.35×10*^−^*^109^; *β* = 2.24), whereas the coding model (ptvraredmg; Methods) was associated with decreased protein levels (*p* = 1.90×10*^−^*^111^; *β* = −1.63; Fig. 4f), underscoring that non-coding (including G4) effects can counteract coding variant effects on protein abundance of the same gene.

We also identified significant non-coding-exclusive (absent in coding models) pQTL associations: 317 in *Baseline non-coding* (26 *cis*, 291 *trans*) and 170 in *G4-Mechanistic non-coding* models (5 *cis* and 165 *trans*). For example, *PTK7* demonstrated a highly significant *cis* association in two G4-disruptive models: promoter (200 bp) (*p* = 4.25 × 10*^−^*^61^; *β* = 1.99) and promoter (1 kb) (*p* = 3.71 × 10*^−^*^60^; *β* = 1.96) (SI Fig. 19a). The same signals were detected in other non-coding models but with reduced effect and significance (G4-modifying promoter (200 bp) model: *p* = 2.61 × 10*^−^*^35^ and *β* = 1.09; G4-modifying promoter (1 kb) model: *p* = 3.87 × 10*^−^*^18^ and *β* = 0.61; all-variant promoter (200 bp) model: *p* = 9.85 × 10*^−^*^15^ and *β* = 0.43; all-variant promoter (1 kb) model: *p* = 1.39 × 10*^−^*^07^ and *β* = 0.21). By contrast, no *PTK7 cis* associations were detected in coding gene-level models even at *p* = 10*^−^*^4^ threshold (PTV model: *p* = 7.13 × 10*^−^*^1^; *β* = 0.26). As a *trans* example, *HYAL1* showed strong associations with ACY1 protein abundance in UTR (template strand) G4-modifying model (*p* = 1.35 × 10*^−^*^36^; *β* = −1.56) and other non-coding models including UTR G4-modifying model (*p* = 1.43 × 10*^−^*^28^; *β* = −1.16), UTR-promoter (1 kb) G4-modifying model (*p* = 5.75 × 10*^−^*^21^; *β* = −0.80), and UTR-promoter (200 bp) G4-modifying model (*p* = 5.75×10*^−^*^21^; *β* = −0.80) but not in any coding models [49] (SI Fig. 19b). Together, these findings suggest a highly specific and robust role for G4-disruptive variants in modulating protein abundance that is captured only when variants are aggregated by mechanism.

Next, we performed non-coding gene-level collapsing analyses for binary and quantitative traits and identified 102 and 166 significant associations (*p <* 10*^−^*^8^; SI Tables 16-17, SI Fig. 17, 20) with FDRs of 3.48% and 1.33%, respectively. Of these associations, 93.1% (binary) and 78.9% (quantitative) are exclusive to collapsing models, with no single non-coding rare variant in the corresponding *regulatory regions* reaching *p <* 10*^−^*^4^ (Fig. 4b). For example, *RILP* (an intracellular effector [62]) was associated with mean diffusivity in the right tapetum on the FA skeleton (*p* = 2.81 × 10*^−^*^9^; *β* = −1.27) only in the UTR-G4 collapsing model, not at the variant level. Like pQTL associations, effect sizes of binary and quantitative associations in the collapsing models followed the same pattern: G4-formative *>* G4-disruptive *>* G4-modifying *>* all-variant (CADD ≥ 5) *>* all-variant models (Fig. 4g-h and SI Table 15).

Of the 102 and 166 significant binary and quantitative associations, 82 (80.4%) and 110 (66.3%) were non-coding-exclusive (SI Fig. 18b), i.e., the same gene-phenotype associations were not observed in coding models. Examples include the *ACTL8* association with bacterial pneumonia (UTR G4-modifying model: *p* = 9.25 × 10*^−^*^9^; OR = 223.33; SI Fig. 21a) and *WNT4* (involved in gonadal development and sex determination [63]) association with reduced alkaline phosphatase levels (UTR G4-disruptive model: *p* = 1.15 × 10*^−^*^22^; *β* = −2.00; SI Fig. 21b). Notably the *WNT4* association is not observed in any coding models, but it aligns with prior reports that reduced *WNT4* expression decreases ALP activity [64]. The remaining 20 (19.6%) and 56 (33.7%) significant binary and quantitative associations were coding-supported (SI Fig. 18b). These findings revealed novel disease and trait associations with non-coding regulatory elements, including G4 structures, that were missed by coding models, thereby expanding our understanding of genetic contributions to human phenotypes. Furthermore, these gene–phenotype associations overlapped substantially across models, consistent with shared regulatory architecture, yet each model also captured a distinct subset of associations, supporting the complementary value of different regulatory annotations (SI Fig. 17).

To be noted, a potential concern is that selecting only a few variants might randomly reduce effect cancellation and inflate signals. However, compared with all *G4-Mechanistic non-coding* tests, significant *G4-Mechanistic non-coding* associations involved a greater average number of variants per test: 1.5× more in the G4-modifying and G4-disruptive models, and roughly 4× more in the G4-formative models (SI Table 12; SI Fig. 14). This indicates that the observed significant associations are likely driven by enrichment of functional G4 variants with concordant effect directions rather than spurious signals arising from randomly reduced variant heterogeneity or effect cancellation. These results show that mechanism-informed collapsing models centered on G4 biology identified novel associations with stronger effects and mitigated limitations inherent to traditional rare variant collapsing approaches.

### Non-coding-augmented PTV collapsing reveals novel associations

PTV collapsing models capture coding loss-of-function effects, but can miss regulatory effects acting through non-coding elements. To obtain a more complete view of gene-level impact, we implemented gene-level collapsing models that incorporated PTVs (from the same gene) into the non-coding models (SI Table 13), hereafter collectively referred to as “PTV + non-coding” (Methods). Across protein abundances and traits, the PTV + non-coding model revealed 80 *cis*-pQTL, 459 *trans*-pQTL, 140 binary, and 164 quantitative associations achieving significance in the PTV + non-coding model, but were not significant in PTV model alone (Fig. 5a, SI Tables 18-20). Of these, 34 *cis*-pQTL, 169 *trans*-pQTL, 88 binary, and 68 quantitative associations were detectable only when PTVs and non-coding variants were analyzed together—not in either model alone (Fig. 5a).

**Fig. 5:**
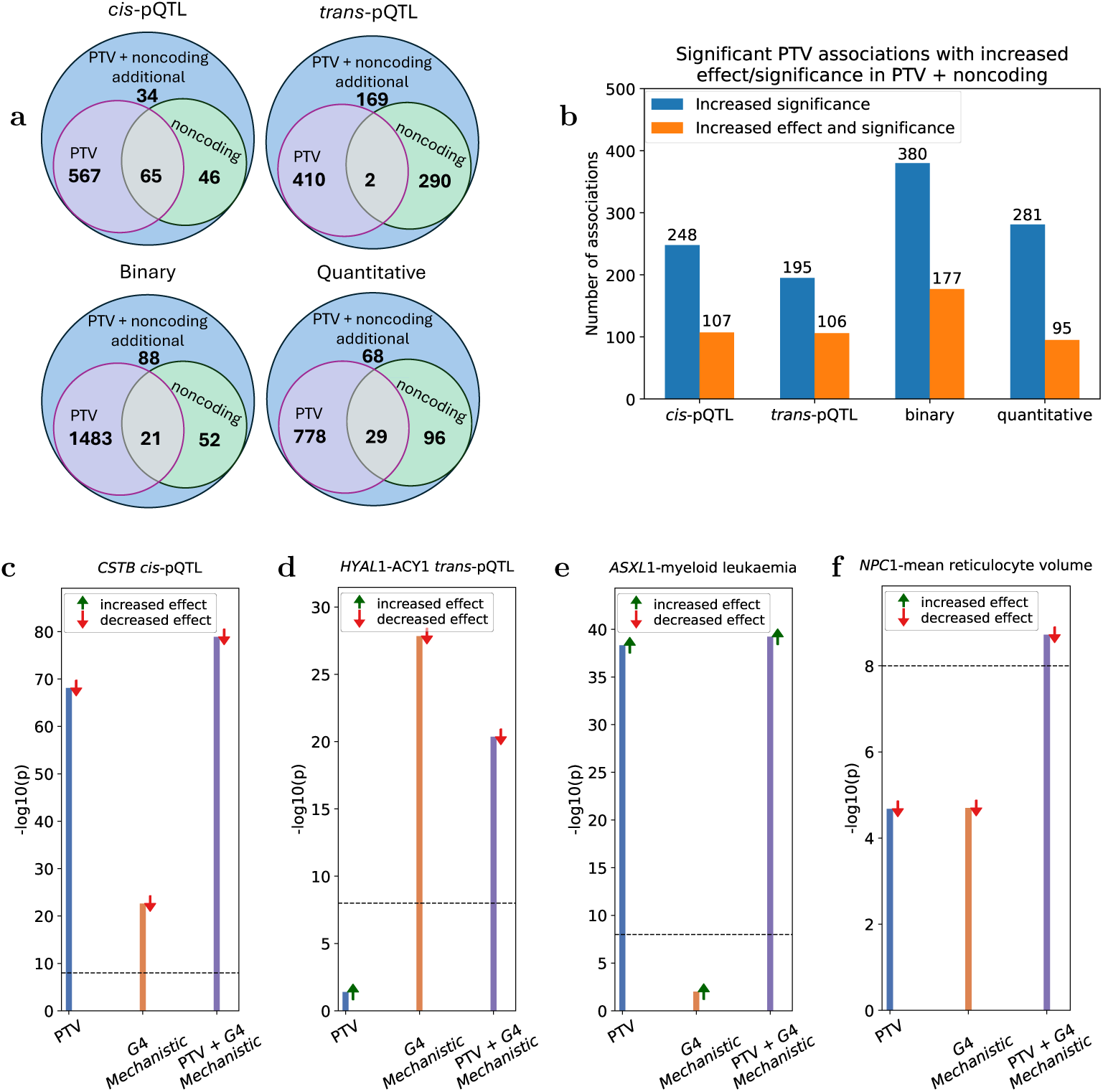
Non-coding-augmented PTV collapsing analyses summary. **(a)** Venn diagram showing additional associations identified by the combined PTV + noncoding model and their overlap with associations detected by the PTV-only and noncoding-only models across phenotypic categories. **(b)** For associations shared between the PTV-only and combined PTV + noncoding models, the bar plot shows the number of associations with greater statistical significance, as well as those with both greater significance and larger effect sizes in the combined model, across phenotypic categories. The *p* values for selected gene-level associations from coding (PTV), non-coding (*Baseline* or *G4-Mechanistic non-coding*), and non-coding-augmented PTV (PTV + *Baseline non-coding* and PTV + *G4-Mechanistic non-coding*) models for **(c)** *cis*-pQTLs, **(d)** *trans*-pQTLs, **(e)** binary traits, and **(f)** quantitative traits. The dashed line marks the study-wide significance threshold (*p <* 10*^−^*^8^). The *Baseline* and *G4-Mechanistic non-coding* collapsing models are detailed in SI Table 13. Coding PTV associations are from refs. [48, 49, 53]. Arrow orientation denotes effect direction: downward red arrow indicates decreased effect; upward green arrow indicates increased effect.

Among associations already significant with the PTV model, the combined model increased significance for 248 *cis*-pQTL, 195 *trans*-pQTL, 380 binary, and 281 quantitative associations (Fig. 5b). Notably, 107 *cis*-pQTL, 106 *trans*-pQTL, 177 binary, and 95 quantitative associations not only became more significant but also demonstrated larger effect sizes in the PTV + non-coding model (Fig. 5b). For example, *cis*-pQTL signals for *CSTB* were robust in both the PTV model (*p* = 7.81 × 10*^−^*^69^; *β* = −1.83) and the non-coding promoter G4-modifying model (*p* = 2.58×10*^−^*^20^; *β* = −0.79), with the combined model (Fig. 5c) further strengthening the signal (*p* = 1.85 × 10*^−^*^79^; *β* = −1.24). In contrast, a *trans*-pQTL association of *HYAL1* with ACY1 protein abundance (Fig. 5d) showed decreased effect size and reduced significance in the combined model (*p* = 4.40×10*^−^*^21^; *β* = −0.91), due to cancellation of effects between PTV model (*p* = 4.01 × 10*^−^*^2^; *β* = 0.51) and UTR G4-modifying model (*p* = 1.43 × 10*^−^*^28^; *β* = −1.16). For disease associations, association between *ASXL1* and myeloid leukaemia (Fig. 5e) became more significant in the combined model (*p* = 6.11×10*^−^*^40^; OR = 1.71) than in either the splice-proximal intron-G4-formative-only model (*p* = 1.01 × 10*^−^*^2^; OR = 1.73) or PTV-only model (*p* = 4.71 × 10*^−^*^39^; OR = 1.72). Another example is the association between *NPC1* and mean reticulocyte volume (Fig. 5f) which was missed by both PTV model (*p* = 2.1 × 10*^−^*^5^; *β* = −0.22) and UTR-promoter (1 kb)

G4-modifying model (*p* = 2.0 × 10*^−^*^5^; *β* = −0.19) but became significant in the combined model (*p* = 1.90 × 10*^−^*^9^; *β* = −0.20). The same association was also observed in another coding model (flexdmg: *p* = 2.43 × 10*^−^*^11^; *β* = −0.08; Methods) [48, 53], supporting the robustness of the signal. These results demonstrate clear complementarity between PTVs and non-coding variants. The combined model captures additive signals when mechanisms align and reveals biologically meaningful cancellation when directions oppose.

### Cross-biobank replication highlights sample-size dependence and robust associations

To assess robustness, we validated UKB association results in the independent Alliance for Genomic Discovery (AGD) cohort comprising 195,457 EUR samples and 12,344 binary traits (Methods; SI Table 21). Using the same gene-level collapsing models (SI Table 13) across *regulatory regions*, we compared AGD (195,457 samples) and UKB (459,449 samples) association results across 4,367 binary phenotypes recorded in both cohorts (SI Table 22). Within these shared phenotypes, AGD yielded 49 significant associations (FDR = 3.03%) and UKB yielded 123 (FDR = 3.48%) (Fig. 6a). 85 (70.2%) out of 121 UKB associations (two tests excluded for *<* 5 cases/carriers in AGD cohort) replicated in AGD dataset at nominal significance (*p <* 0.05) (Fig. 6b; SI Fig. 22a). The replication rate for *G4-Mechanistic non-coding* model associations was 50%. We additionally performed variant-level PheWAS for all variants located in promoter (200 bp) in both biobanks for the same 4,367 binary phenotypes (SI Table 23). UKB and AGD identified 24 and 47 significant variant–phenotype associations, respectively (Fig. 6c). 8 (42.9%) out of 17 UKB associations (seven tests excluded due to *<* 5 cases/carriers in AGD cohort) replicated in the AGD dataset at nominal significance (*p <* 0.05) (Fig. 6d; SI Fig. 22b).

**Fig. 6:**
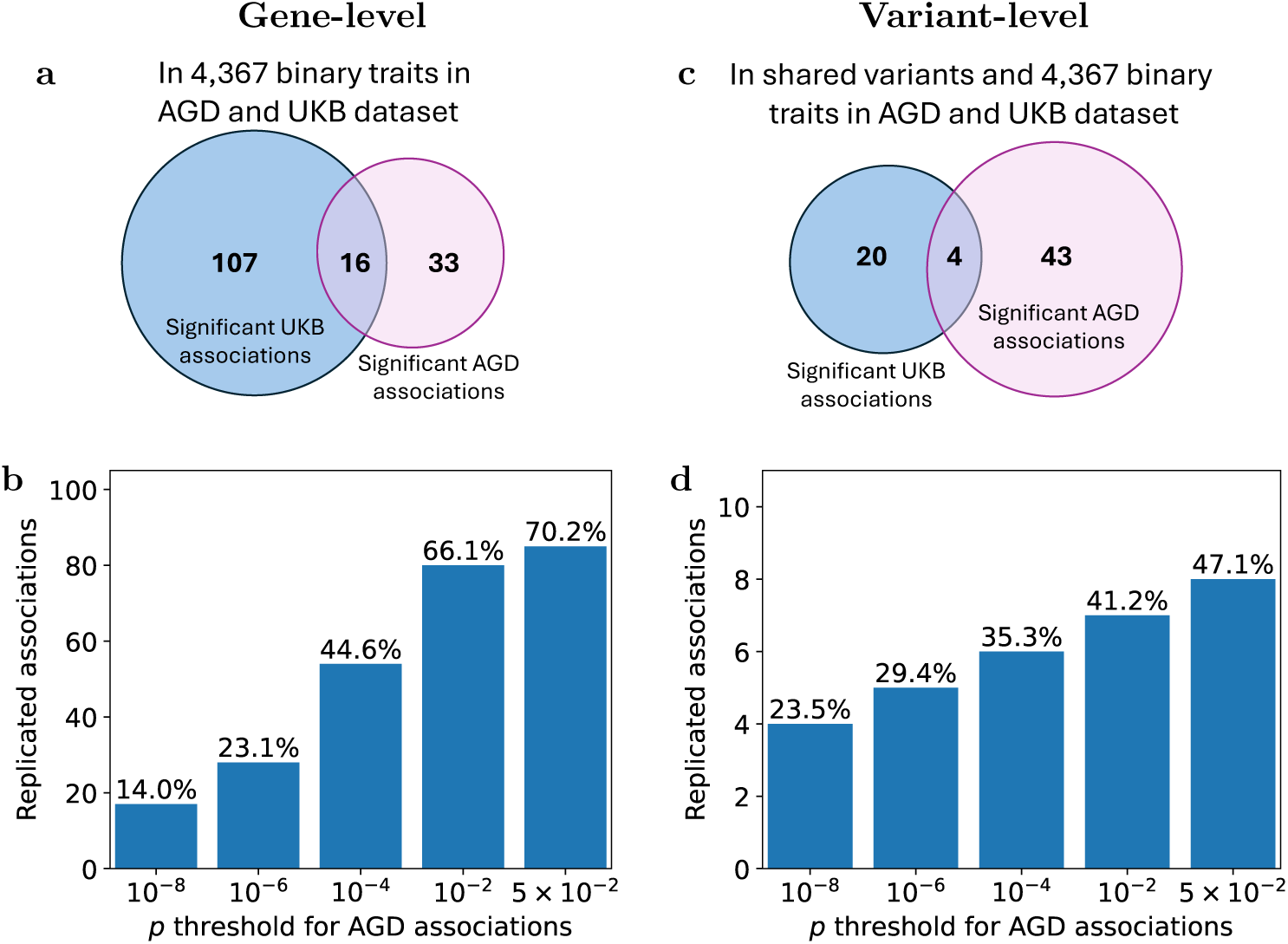
Cross-cohort replication of regulatory associations in AGD and UKB. **(a)** Venn diagram of significant gene-level associations from non-coding *regulatory regions* collapsing models across 4,367 binary traits in AGD (n=195K) and UKB (n=459K) datasets. **(b)** Count and percentage of significant binary associations (*p <* 10*^−^*^8^) in the UKB data replicated (with directional concordance) in the independent AGD dataset at various *p*-value threshold for gene-level analyses. **(c)** Venn diagram of significant variant-level associations in promoter (200 bp) across 4,367 binary traits and genotypes (variants) present in both in AGD and UKB. **(d)** Count and percentage of significant binary associations (*p <* 10*^−^*^8^) in the UKB data replicated (with directional concordance) in the independent AGD dataset at various *p*-value threshold for promoter (200 bp) variant-level analyses.

The replication results provide evidence for the robustness of gene-trait associations across independent cohorts, though differences in cohort size (Methods), phenotype definitions and population characteristics (AGD clinically ascertained vs UKB population-based) may explain the partial overlap.

## Discussion

This study systematically maps non-coding PQ-modifying variants across key *regulatory regions* genome-wide and assesses their functional impact through PheWAS in 459,449 UK Biobank European participants. These variants are widespread across the genome, exert bidirectional functional effects, and often act independently of coding variation in shaping human molecular and phenotypic diversity. By explicitly modelling G4-disruptive, G4-formative, and G4-modulatory variants as well as aggregating them in mechanism-informed collapsing tests, we uncover associations with larger effects and improved sensitivity relative to conventional rare-variant approaches. These results reposition G4s from biophysical features to practical, genome-wide mediators of regulatory variation, providing a mechanistic lens for interpreting non-coding genetic variation effects on human traits.

The balanced distribution of significantly associated G4 variants between the template and non-template strands suggests that observed G4 functional effects in this study largely involve DNA-mediated mechanisms. However, it is important to emphasize that our study focuses on putative G4s identified in DNA sequences, not spliced transcripts, and many of the significant UTR G4 variants fall in intronic regions. Our study is therefore not designed to study potential functional G4 relationships that act at the RNA level.

A central advance of this work is a mechanism-informed collapsing framework tailored to G4 biology. Traditional non-coding collapsing models (for example, all rare variants, or filters based on scores like CADD) assume largely unidirectional effects and are vulnerable to effect cancellation when non-coding variants act in both positive and negative directions. In contrast, *G4-Mechanistic non-coding* collapsing selects variants by predicted structural impact (formative, disruptive, modulatory), thereby enriching for functionally coherent variants and reducing cancellation. Across *regulatory regions*, gene-level *G4-Mechanistic non-coding* models produced substantially stronger effects than *Baseline non-coding* models, with a consistent patter in effect sizes: G4-formative *>* G4-disruptive *>* G4-modifying *>* baseline burdens. Non-coding-exclusive associations underscore distinct regulatory contributions alongside protein-altering effects. Replication with directional concordance in an independent AGD cohort (70.2% at the gene level and 47.1% at the variant level), despite differences in cohort size and ascertainment, supports robustness and generalizability. Integrating non-coding variants with PTV in a combined model (“PTV + non-coding”) further improved association discovery, revealing 843 significant associations undetected by the PTV-only model alone and 359 missed by both PTV-only and non-coding-only models, indicating complementary information across mechanisms. In some genes, signals strengthened when mechanisms aligned; in others, opposing directions revealed biologically meaningful cancellation. These results would be overlooked in single-model analyses. Combining coding and non-coding variants yielded a more comprehensive view of gene-level impact on phenotypes.

A few limitations should be acknowledged. First, this analysis focused on canonical G4s, excluding non-canonical G4 structures such as bulged G4s [13], which may add regulatory complexity not captured here. Second, G4 formation is likely cell-type specific [65], but our PheWAS framework does not yet incorporate tissue- or cell-type–resolved information and is restricted to 2,941 plasma proteins, potentially obscuring context-specific effects. Third, annotated G4 regions frequently overlap with other regulatory pathways, complicating attribution specifically to G4 formation or disruption. Our functional categorization of G4 variants (disruptive, formative, modulatory) is predictive and requires biochemical and cellular assays to substantiate these assignments. Finally, the observed associations between putative G4-modifying variants and phenotypes are correlative, and causal inference will require experimental validation.

Overall, the combination of genome-scale mapping, mechanism-informed collapsing models, and phenome-wide association analyses established G4s as a key axis of non-coding regulation with immediate implications for functional genomics and therapeutic modulation. Our analysis prioritized candidate G4 loci that can be targeted with small molecules, modulators of G4-binding proteins, antisense or synthetic oligonucleotides, and CRISPR-based tools to link genotype G4 structure and gene expression. Looking ahead, extending this framework to other secondary structures (such as hairpins, Z-DNA, and inverted repeats) across non-coding regions should further elucidate the non-coding genome and accelerate translation to human health.

## Methods

### UKB resource

The protocols for the UKB are overseen by the UKB Ethics Advisory Committee. More information can be found at https://www.ukbiobank.ac.uk/ethics and https://www.ukbiobank.ac.uk/wp-content/uploads/2025/01/Ethics-and-governance-framework.pdf. All participants provided informed consent. The study protocol and operational processes were reviewed and approved by the Northwest Research Ethics Committee (REC reference 06/MRE08/65). Data access and analyses for the current work were conducted under UK Biobank applications 24898 and 68574.

#### WGS and sample selection

Whole-genome sequencing (WGS) was performed for 490,560 UK Biobank participants through a public–private collaboration that included deCODE Genetics, the Wellcome Trust Sanger Institute, AstraZeneca, Amgen, GlaxoSmithKline, Johnson & Johnson, UK Research and Innovation, and the UK Biobank. The sequencing and quality-control procedures are detailed elsewhere [53]. In brief, genomic DNA was sequenced on Illumina NovaSeq6000 platforms using paired-end reads of 2 × 151 bases, with a mean depth of 32.5×. The DRAGEN 3.7.8 pipeline was used for variant calling as described in ref. [53], including its running mode, parameterization, and computational requirements.

A series of sample-level filters were applied prior to analysis: (1) exclusion of participants who withdrew consent; (2) removal of samples with less than 2% concordance relative to whole-exome sequencing and genotyping array data; (3) elimination of suspected duplicate samples associated with multiple reported birth events; (4) exclusion of samples exhibiting evidence of contamination, defined as *verifybamid freemix* ≤ 0.04 as assessed with VerifyBAMID; (5) removal of samples with insufficient CCDS coverage, i.e., fewer than 94.5% of CCDS r22 bases covered at ≥ 10×; and (6) exclusion of coverage uniformity outliers, defined by a uniformity metric *>* 0.433 (exceeding six times the interquartile range). Population labels were assigned using *peddy* [66] in conjunction with 1000 Genomes reference data, requiring *peddy prob* ≥ 0.9. European-ancestry samples were further partitioned with the gnomAD classifier [8] into non-Finnish European (NFE) and Ashkenazi Jewish groups. Additional quality control within the NFE subset used principal components derived by *peddy* ; samples lying more than four standard deviations from the mean across the first four principal components were excluded.

#### Olink proteogenomics study cohort

The Olink proteomics cohort has been described extensively in prior reports [6, 49, 67]. For this analysis, we examined 49,736 samples with matched whole-genome sequencing data. In total, measurements for 2,941 plasma proteins were included (listed in SI Table 3).

#### Phenotypes

We analyzed two broad phenotype classes—binary and quantitative—drawn from the April 2023 UK Biobank release, accessed on 28 June 2024 under applications 68601 and 65851. Phenotype extraction and harmonization were performed with our previously described PEACOK package https://github.com/astrazeneca-cgr-publications/PEACOK, as detailed in prior work [48]. In total, the study encompassed 13,321 binary traits and 1,682 quantitative traits (listed in SI Table 3).

For binary outcomes, control selection followed our established procedure [48]: when the proportion of female cases differed significantly from that of available female controls (two-sided Fisher’s exact test, *p <* 0.05), controls were sex-matched to cases. This rule also covered sex-specific traits, for which all controls were, by design, of the same sex as cases [48].

#### Relatedness pruning

Relatedness pruning was performed with the *ukb gen samples to remove()* function from the *ukbtools* R package (v0.11.3) [68]. We targeted a sample set in which no pair of individuals had a kinship coefficient greater than 0.0884, corresponding approximately to third-degree relatives. For each related pair, the procedure discarded the individual with the larger number of relatives above the threshold, yielding a maximal subset that minimizes pairwise relatedness.

#### QC filters

In this work, we utilised 459,449 unrelated NFE participants from the UK Biobank dataset. Details on the initial processing of the genomic data are in ref. [53] and briefed in the previous subsection. For this analysis, we removed any variant failing any of the following criteria: Genotype quality ≥ 20, Depth ≥ 10, missing call rate *<* 5%, Fisher Strand Bias (FS) ≥60 for SNVs and ≥200 for indels, DRAGEN variant status = PASS, and root-mean-square mapping quality (MQ) *>*=30. This work includes only short variants, specifically single-nucleotide variants and small insertions and deletions (≤ 50 bp).

After applying stringent variant QC filters, we extracted 32,269,210 single-nucleotide variants and 4,545,980 small insertions and deletions (≤ 50 bp) located within *regulatory regions* from 459,449 UKB NFE ancestry participants. The count of G4 variants are provided in SI Table 2.

### Alliance for Genomic Discovery cohort

We conducted replication analyses using data from the Alliance for Genomic Discovery (AGD), also referred to as the NashBio cohort (www.nashbio.com). This resource includes WGS data from approximately 250,000 participants (likely patients) across the United States collected at the Vanderbilt University Medical Center’s BioVU® biobank and representation across diverse ancestral backgrounds. The BioVU® biobank, administered by Vanderbilt University, provided de-identified DNA samples extracted from discarded clinical blood samples collected from individuals who provided consent or assent, along with corresponding de-identified electronic health records. Whole-genome sequencing was carried out by deCODE genetics using Illumina sequencing platforms. We used the same variant filtering protocol for AGD data as described above for UKB, with the exception that the missing-call rate threshold (*<* 5%) was not applied. The AGD cohort includes 12,344 linked binary traits, of which 4,367 are shared with the UKB binary phenotypes. A complete list of binary traits used in this study is provided in SI Table 21.

Association analyses were performed for individuals of EUR ancestry only. For the gene-level analysis, we tested non-coding collapsing models across *regulatory regions* for 12,344 binary traits in two sets of AGD cohort: 117,016 (subset cohort) and 195,457 (full cohort) EUR samples, to evaluate the effect of cohort size on association results. All non-coding models described in SI Table 13 were evaluated. Significant associations identified in the AGD data are provided in SI Table 24 for the subset (117K) cohort and in SI Table 22 for the full (195K) cohort. To estimate the significance threshold, we applied an *n*-of-1 permutation framework, analogous to that used for the UKB data, to estimate the false discovery rate (FDR). To enable systematic comparison with UKB non-coding association results, the significance threshold for AGD associations was set to 3 × 10*^−^*^9^ (FDR = 3.03%), comparable to the FDR of gene-level binary associations in the UKB data (FDR = 3.48%).

For the variant-level analysis, we restricted to variants present in at least five participants in promoter (200 bp), and tested associations with AGD binary phenotypes shared among UKB, as well as in the full AGD cohort. The significant associations are provided in SI Table 23.

We conducted validation analyses in Alliance for Genomic Discovery (AGD) data (see Methods). We first performed gene-level association analyses (SI Table 13) across *regulatory regions* for 12,344 linked binary traits (SI Table 21) in two sets of AGD cohorts: a full cohort (195,457 EUR samples) and its subset cohort (117,016 EUR samples). The subset cohort identified 23 significant associations (SI Table 24), whereas the full cohort revealed 66 significant associations (SI Table 22), with 15 associations achieving significance in both datasets (SI Fig. 23). A 1.67-fold increase in sample size yielded a 2.87-fold increase in significant discoveries, demonstrating that sample size critically influences the detection of association signals. These results highlight how larger cohorts enable discovery of more associations and underscore the value of continued biobank growth.

### G4s and G4-modifying variants

Putative G4s were identified by searching for the motif {G_3+_L_1-12_}_3+_G_3+_ [11] where G denotes Guanine and L is loop of 1-12 nucleotides of any base. All identified putative G4s in *regulatory regions* are provided in SI Table 1. Experimentally validated G4s were obtained from high-throughput G4-seq data [13].

G4 variants are classified as formative, disruptive, or modulatory based on their predicted ability to form or disrupt or modulate a G4 structure in the alternate allele. For each variant, we analyzed a 50 base pair region flanking the variant position both upstream and downstream. A variant is classified as disruptive if a G4 consensus sequence in the reference, involving the variant position, disappears in the alternate. It is considered formative if the alternate introduces a G4 consensus sequence absent in the reference. Variants that extend or shorten G4s while leaving consensus sequence unchanged (for example, changes that increase or decrease the number of G-tracts, or alter the length of any loop or G-tract while still satisfying the consensus sequence) are labeled as modulatory variants. The distribution of G4-modifying variants across *regulatory regions* is provided in SI Table 2.

### Regulatory region coordinates

We used the reference genome build GRCh38. We explored variants in five key non-coding *regulatory regions* of the human genome including promoters, 3*′*UTRs, 5*′*UTRs, splice-proximal introns, and CTCF binding sites. The coordinates for these non-coding *regulatory regions* were taken from GENCODE version 43. We analysed autosomes only and removed non-coding fragments overlapping coding sequences. CTCF binding sites were identified using Find Individual Motif Occurrences (FIMO) [69], considering only matches with a significance threshold of *p <* 10*^−^*^7^ which led to 8,830 CTCF binding sites across the autosomes. For the analysis, we considered a 200 bp window centered on each predicted 18 bp CTCF binding site. For splice-proximal introns, we took 100 bp into the intron from each annotated exon-intron boundary, respecting transcript strand. No bases from exonic sequence were included.

### Mutability

We aimed to compare the mutability of G4 regions with their underlying (host or encompassing) regulatory regions, for example, comparing the mutability of 3*′*UTR G4s relative to the overall 3*′*UTR region. Mutability of a region is computed as the average burden of rare variants (MAF *<* 0.1%) per base pair within that region and normalised by sample size:

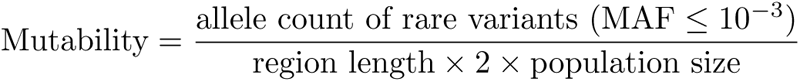

Note that, by focusing on recent, minimally selected variants, this analysis primarily reflects local mutation rates rather than functional importance.

### Variant-level association analyses

We performed association tests for variants observed in at least five participants within the 459,449 unrelated NFE UKB genomes or 46,327 unrelated NFE UKB genomes in case of proteomic association analysis. Variant-level association studies were performed for all variants in promoters (200 bp) and CTCF binding sites and only for putative G4-modifying variants in 3*′*UTRs, 5*′*UTRs, and splice-proximal introns. For binary phenotypes, we calculated variant-level *p*-values using a two-sided Fisher’s exact test under a dominant genetic model (carriers: AB/AA vs non-carriers: BB, with A as the alternate and B as the reference allele). For quantitative phenotypes, association testing was performed with linear regression models that included age, sex, and (age × sex) as covariates.

All significant variant-level associations (*p <* 10*^−^*^8^) observed in UKB data are provided in SI Tables 4, 7, and 8. We also compared our results with prior association studies of coding variants reported by Wang et al. [48], Dhindsa et al. [49], and the UK Whole Genome Consortium [53]. Variants associated with the protein abundance of a gene were classified as cis-pQTLs if they were located within the genomic span of the same gene (including its regulatory regions) and as trans-pQTLs if they were located outside the genomic span of that gene. An analogous definition applies to gene-level collapsing analyses.

### Gene-level collapsing association analyses

We performed gene-level analyses by collapsing (combining multiple variants within a region into a single participant-level score [48, 50]) all rare variants (MAF *<* 0.1%) in gene-associated non-coding regulatory elements, including promoter (200 bp), promoter (1 kb), splice-proximal introns, UTRs and combined regions including UTR-promoter (200 bp) and UTR-promoter (1 kb). Then in other collapsing models, rare variants were further prioritised using CADD (Combined Annotation-Dependent Depletion [70]) scores and predicted impact on G4-structure for G4-modifying variants. This way, we classified all collapsing models into two categories: *Baseline non-coding* and *G4-Mechanistic non-coding*. The *Baseline non-coding* models include two collapsing strategies that collapse: (a) all rare variants and (b) rare variants with a CADD score ≥ 5. The *G4-Mechanistic non-coding* models include three collapsing strategies that collapse: (c) G4-modifying rare variants, (d) G4-disruptive rare variants, and (e) G4-formative rare variants. Each strategy was applied separately to promoters (1 kb), promoters (200 bp), UTRs, splice-proximal introns, UTR-promoters (1 kb), and UTR-promoters (200 bp), resulting in 30 collapsing models. In addition, for splice-proximal introns and UTRs, analogous template and non-template *G4-Mechanistic non-coding* models were constructed using G4 variants restricted to the template or non-template strands, yielding six additional models. In total, 36 collapsing models were evaluated based on five distinct collapsing strategies. These models are also summarised in SI Table 13. All collapsing models followed a dominant scheme, where carriers of at least one qualifying variant in non-coding regulatory regions were compared with non-carriers. Similar to variant-level association analysis, *p*-values for collapsing analysis were generated by using Fisher’s exact test for binary traits and linear regression for quantitative traits using age, sex and age × sex as covariates.

All gene-level significant associations (*p <* 10*^−^*^8^) observed in UKB dta are provided in SI Tables 14, 16, and 17. Finally, we compared our findings with prior UK Biobank association studies of coding gene-level models [48, 49, 53], including syn (synonymous negative control), ptv (protein-truncating), ptv5pct (protein-truncating; ≤5% MAF), UR (ultra-rare damaging), Urmtr (ultra-rare damaging, Missense Tolerance Ratio [MTR]–informed), raredmg (rare damaging), raredmgmtr (rare damaging, MTR-informed), flexdmg (flexible MAF, damaging non-synonymous), flexnonsynmtr (flexible MAF, non-synonymous, MTR-informed), ptvraredmg (combined PTV or rare damaging), and rec (non-synonymous recessive). Additional methodological details on gene-level coding models are provided in the supplementary information of ref. [53].

### Non-coding-augmented PTV models

We extended our analysis by integrating PTVs into the non-coding models to create combined burden models, collectively referred to as “PTV + non-coding” models. Specifically, PTVs were added individually to each of the five collapsing strategies across promoter (200 bp), promoter (1 kb), splice-proximal intron, UTR, UTR-promoter (200 bp), and UTR-promoter (1 kb) regions, as described in SI Table 13. Thus, every non-coding model in SI Table 13 was evaluated separately in combination with the corresponding PTV model. For each gene, carriers of either class (PTVs or non-coding variants) were collapsed and analysed using the same approach as in the original PTV-only or non-coding-only models.

All significant associations (*p <* 10*^−^*^8^) for PTV + non-coding models are provided in SI Tables 18-20.

### Assessment of genomic inflation factor, *λ*

Genomic inflation factor (*λ*) was calculated as the ratio of the median observed test statistic to the median test statistic obtained from n-of-1 permutation [48, 71] derived null *p*-values. *λ* statistics and distribution plots for variant-level and gene-level models across phenotype categories are in SI Table 25 and SI Fig. 24. The median genomic inflation (*λ*) was 1.033 for binary traits (Fisher’s exact test) and 1.002 and 1.014 for quantitative traits and plasma protein levels (linear regression) (SI Fig. 24 and SI Table 25) for variant-level models. *λ* was 0.995 for binary traits (Fisher’s exact test) and 1.016 and 1.005 for quantitative traits and plasma protein levels (linear regression) (SI Fig. 24 and SI Table 25) for gene-level collapsing models. Taken together, these results indicate that our tests show minimal susceptibility to systematic bias or inflation.

### Defining a unified study-wide significance threshold

We used a study-wide significance threshold of *p <* 1×10*^−^*^8^. To estimate this threshold, we applied n-of-1 permutation as in ref. [48, 71] and generated an empirical null by permuting phenotype labels (case–control or quantitative) once per trait, while preserving the original participant-genotype assignments across all variant- and gene-level tests. The false discovery rate (FDR) was then estimated as the proportion of associations exceeding the significance threshold in the permuted run relative to the corresponding number in the original analysis. We obtained FDRs of 0.022%, 0.005%, and 0.082% for variant-level protein abundances, quantitative traits, and binary traits association analyses, respectively, and 1.46%, 1.33%, and 3.48% for gene-level protein abundances, quantitative traits, and binary traits association analyses, respectively (see SI Table 26).

We used AGD data for replication analyses and applied an analogous n-of-1 permutation framework to estimate FDR. To enable systematic comparison with UKB non-coding association results, the significance threshold for AGD associations was set at 3 × 10*^−^*^9^ (FDR = 3.03%), matching the FDR of gene-level binary associations in the UKB data (FDR = 3.48%).

### Linkage disequilibrium analyses for significantly associated non-coding variants

We performed linkage disequilibrium (LD) analyses for all significantly (*p <* 10*^−^*^8^) associated non-coding variants identified in the variant-level PheWAS of UK Biobank samples. To compare these associations with known genetic signals, we retrieved all genome-wide significant (*p <* 10*^−^*^8^) associations from the NHGRI-EBI GWAS Catalog (accessed 22 March 2026).

LD analyses were restricted to single-nucleotide GWAS variants reported in EUR ancestry studies and located within a ±500 kb window of each significantly associated non-coding variant. Using PLINK v2.00 [72], we computed pairwise LD between each GWAS variant and the corresponding non-coding variant. Stratified summary results for rare and common significant non-coding variants are provided in SI Table 10 and complete results are provided in SI Table 11.

A non-coding variant was classified as being in LD with a GWAS variant if R^2^ ≥ 0.2 and the GWAS variant was also associated with the same phenotype. Pheno-type similarity was assessed using Levenshtein distance [73], implemented in Python with the fuzzywuzzy 0.18.0 library using the partial ratio function. Phenotypes were considered similar if the resulting similarity score was *>* 0.8.

## Data availability

Annotations for putative G4s studied in this work are provided in SI Table 1. UKB phenotypes are listed in SI Table 3 and all significant associations in UKB data set for non-coding models are provided in SI Tables 4, 7, 8, 14, 16, and 17. All significant associations in UKB data for PTV + non-coding models are in SI Tables 18-20. All phenotypes and significant associations in the AGD data are provided in SI Tables 21-23.

## Code availability

PheWAS association tests were conducted using PEACOK. The PEACOK package is a custom framework and is available on GitHub (github.com/astrazeneca-cgr-publications/PEACOK). All code used in the work is described in the Methods and the Reporting Summary. Unless otherwise specified in the Methods, all analyses and plotting were performed using Python 3.9.5.

## Acknowledgements

This research was funded by AstraZeneca. R.S. is a fellow of the AstraZeneca Post-doctoral Research Programme. We are indebted to the volunteers and researchers of UKB and AGD; their contributions made this research possible. UKB cohort: This research was conducted using the UK Biobank Resource under Application Numbers 68601 and 65851. AGD cohort: The samples and structured clinical data were obtained from NashBio, derived from Vanderbilt University Medical Center’s BioVU® biobank. BioVU® is supported by institutional funding, private agencies, and federal grants (including NIH funded Shared Instrumentation Grant S10OD017985, S10RR025141, and S10OD025092; as well as CTSA grants UL1TR002243, UL1TR000445, and UL1RR024975). We thank BioVU participants for their generous contributions. Sequencing of 280,000 BioVU® WGS individuals was funded by the Alliance for Genomic Discovery (NashBio, Illumina, and industry partners Alnylam, Amgen, Abb-Vie, AstraZeneca, Bayer, BMS, GSK, Merck Sharp & Dohme, and Novo Nordisk). DNA sequencing was performed by deCODE genetics using Illumina technology. Individual-level raw data is not available due to dataset nature and commercial limitations.

## Author contributions

R.S., S.P., D.G., Q.W., and X.Z.Z. designed the study. R.S. performed the initial data processing, with support from M.K., K.K., I.G.S., S.A., and X.Z.Z. R.S., F.H., X.L., and R.C. conducted the PheWAS analyses with input from X.Z.Z., Q.W., and A.M.M. R.S. and X.Z.Z. analysed the results and drafted the manuscript, with input from all authors. All authors reviewed and approved the final manuscript.

## Ethics declarations

R.S., F.H., X.L., R.C., K.K., S.A., M.K., S.W., S.M., D.V., R.S.D., O.S.B., Q.W., S.P., and X.Z.Z. were employees, contractors or paid consultants of AstraZeneca during this study and may hold AstraZeneca stock or stock options. The remaining authors declare no competing interests.

