## Supplementary Information for "The Impact of Non-coding G-quadruplex Variants on Human Traits and Disease Susceptibility"

#### Supplementary Tables

**SI Table 1**: Table (provided separately as an Excel file) listing the G4 coordinates analyzed across *regulatory regions*, including 3′UTRs, 5′UTRs, CTCF binding sites, promoters (200 bp), promoters (1 kb), and splice-proximal intron (100 bp). Variant-level analyses were performed only for 3′UTRs, 5′UTRs, CTCF binding sites, promoters (200 bp), and splice-proximal introns (100 bp). This file also contains the list of G4s for promoters (1 kb), which were included only in the gene-level analysis.

| | **G4-disruptive** | **G4-formative** | **G4-modulatory** |
|---|---|---|---|
| 3′UTRs | 49401 | 22456 | 210432 |
| 5′UTRs | 41831 | 20591 | 231304 |
| CTCF binding sites | 6993 | 4360 | 36470 |
| promoters (1 kb) | 121447 | 60184 | 808865 |
| promoters (200 bp) | 52357 | 25077 | 424168 |
| splice-proximal introns (100 bp) | 115617 | 41174 | 653055 |

**SI Table 2**: Number of G4-modifying variants across *regulatory regions*.

**SI Table 3**: Table (provided separately as an Excel file) listing UKB phenotypes considered in this study.

**SI Table 4**: Table (provided separately as an Excel file) listing the significant associations ($p < 10^{-8}$) identified from variant-level PheWAS of protein abundances in the UKB dataset.

| **region** | ***cis*-pQTL associations** | ***cis*-pQTLs** | ***trans*-pQTL associations** | ***trans*-pQTLs** |
|---|---|---|---|---|
| | | count for common G4-modifying variants | | |
| promoter (200 bp) | 307 (30, 13, 264) | 306 (30, 13, 263) | 3536 (448, 505, 2583) | 1174 (98, 55, 1021) |
| 3′UTRs | 140 (13, 16, 111) | 140 (13, 16, 111) | 1741 (343, 117, 1281) | 870 (135, 69, 666) |
| 5′UTRs | 70 (6, 4, 60) | 70 (6, 4, 60) | 1123 (184, 140, 799) | 425 (60, 38, 327) |
| splice-proximal introns | 584 (63, 31, 490) | 581 (63, 31, 487) | 7957 (739, 335, 6883) | 2740 (330, 156, 2254) |
| CTCF binding sites | 0 (0, 0, 0) | 0 (0, 0, 0) | 308 (33, 16, 259) | 115 (14, 9, 92) |
| | | count for rare G4-modifying variants | | |
| promoter (200 bp) | 32 (4, 0, 28) | 32 (4, 0, 28) | 78 (7, 6, 65) | 75 (7, 6, 62) |
| 3′UTRs | 7 (1, 0, 6) | 7 (1, 0, 6) | 79 (19, 8, 52) | 65 (15, 6, 44) |
| 5′UTRs | 7 (2, 0, 5) | 7 (2, 0, 5) | 34 (8, 1, 25) | 32 (7, 1, 24) |
| splice-proximal introns | 28 (3, 1, 24) | 28 (3, 1, 24) | 206 (32, 9, 165) | 179 (32, 7, 140) |
| CTCF binding sites | 0 (0, 0, 0) | 0 (0, 0, 0) | 11 (2, 1, 8) | 11 (2, 1, 8) |

**SI Table 5**: Counts of pQTL associations and distinct pQTLs for G4-modifying variants across *regulatory regions*, stratified by allele frequency (common vs. rare; MAF $< 0.1\%$). Counts are further stratified by G4-modifying variant types: G4-disruptive, G4-formative, and G4-modulatory.

| Variants | Type | % positive cis-pQTL | % positive trans-pQTL |
|---|---|---|---|
| Rare | PTV | 3.10% | 56.12% |
| | Non-coding | 51.27% | 35.9% |
| Common | PTV | 16.48% | 54.28% |
| | Non-coding | 46.93% | 55.41% |

**SI Table 6**: Percentage of positive *cis*- and *trans*-pQTL associations ($p < 10^{-8}$) stratified by variant type (PTV vs. non-coding) and allele frequency (common vs. rare; MAF $< 0.1\%$). For non-coding variants, the reported values represent the mean across various non-coding *regulatory regions* analysed in this study. The percentage of positive pQTL associations across non-coding *regulatory regions* is shown in SI Fig. 6.

**SI Table 7**: Table (provided separately as an Excel file) listing the significant associations ($p < 10^{-8}$) identified from variant-level PheWAS of binary traits in the UKB dataset.

**SI Table 8**: Table (provided separately as an Excel file) listing the significant associations ($p < 10^{-8}$) identified from variant-level PheWAS of quantitative traits in the UKB dataset.

| | |
|---|---|
| **Variant** | 11-835996-C-T (G4-modulatory variant on template strand of *CD151* splice-proximal intron) |
| **Coords** | chr11-835974:836076 |
| **Ref.** | CCGCG CCTGG **CCC**TG TGT**CC** **CC**TCC TATCC GTCT**C** **CC**AGT CAGGG G**CCC**G GTGCT GTGG**C** **CCC**GC TGA**CC** **CCTCC** **CC**TGC CTCCT CAG**CC** **CC**AGG ATGGG TG |
| **Alt.** | CCGCG CCTGG **CCC**TG TGT**CC** **C**TTCC TATCC GTCT**C** **CC**AGT CAGGG G**CCC**G GTGCT GTGG**C** **CCC**GC TGA**CC** **CCTCC** **CC**TGC CTCCT CAG**CC** **CC**AGG ATGGG TG |
| **Variant** | 11-835982-T-C (G4-modulatory variant on template strand of *CD151* splice-proximal intron) |
| **Coords** | chr11-835974:836076 |
| **Ref.** | CCGCG CCTGG **CCC**TG TGT**CC** **CC**TCC TATCC GTCT**C** **CC**AGT CAGGG G**CCC**G GTGCT GTGG**C** **CCC**GC TGA**CC** **CCTCC** **CC**TGC CTCCT CAG**CC** **CC**AGG ATGGG TG |
| **Alt.** | CCGCG **CCC**GG **CCC**TG TGT**CC** **CC**TCC TATCC GTCT**C** **CC**AGT CAGGG G**CCC**G GTGCT GTGG**C** **CCC**GC TGA**CC** **CCTCC** **CC**TGC CTCCT CAG**CC** **CC**AGG ATGGG TG |
| **Variant** | 11-836038-G-A (G4-modulatory variant on template strand of *CD151* splice-proximal intron) |
| **Coords** | chr11-835974:836076 |
| **Ref.** | CCGCG CCTGG **CCC**TG TGT**CC** **CC**TCC TATCC GTCT**C** **CC**AGT CAGGG G**CCC**G GTGCT GTGG**C** **CCC**GC TGA**CC** **CCTCC** **CC**TGC CTCCT CAG**CC** **CC**AGG ATGGG TG |
| **Alt.** | CCGCG CCTGG **CCC**TG TGT**CC** **CC**TCC TATCC GTCT**C** **CC**AGT CAGGG G**CCC**G GTGCT GTGG**C** **CCC**AC TGA**CC** **CCTCC** **CC**TGC CTCCT CAG**CC** **CC**AGG ATGGG TG |
| **Variant** | 11-837913-C-T (G4-disruptive variant on template strand of *CD151* splice-proximal intron) |
| **Coords** | chr11-837891:837947 |
| **Ref.** | GTGGG AGGTG **CCCCC** TGGG**C** **CC**GCC TTCAA CA**CCC** ATCCG CG**CCC** **C**GCAG GGCGG C |
| **Alt.** | GTGGG AGGTG *CCCCC* TGGGC CTGCC TTCAA CA*CCC* ATCCG CG*CCC* *C*GCAG GGCGG C |
| **Variant** | 22-24603137-C-CGCCCG (G4-modulatory variant on template strand of *GGT1* promoter) |
| **Coords** | chr22-24603106:24603147 |
| **Ref.** | TTTAA AGCCT **CCCCT** **CCCCC** **CGCCC** **CGCCC** **CC**AGG CCACT AG |
| **Alt.** | TTTAA AGCCT **CCCCT** **CCCCC** **CGCCC** **CGCCC** CGCCC GCAGG CCACT AG |
| **Variant** | 22-24603131-C-G (G4-disruptive variant on template strand of *GGT1* promoter) |
| **Coords** | chr22-24603106:24603148 |
| **Ref.** | TTTAA AGCCT **CCCCT** **CCCCC** **CGCCC** **CGCCC** **CC**AGG CCACT AG |
| **Alt.** | TTTAA AGCCT *CCCCT* *CCCCC* *C*GCCG CG*CCC* *CC*AGG CCACT AG |
| **Variant** | 17-7179137-T-C (G4-modulatory variant on non-template strand of *ASGR1* 5′UTR (intronic)) |
| **Coords** | chr17-7179091:7179192 |
| **Ref.** | TCTGA TGTTT **CCCAC** **CC**AGC A**CCCC** **CC**AAC ACA**CC** **C**TGGG TT**CCC** TCCGC A**CCCC** TGCA**C** **CCCC**A GCCAG CCT**CC** **C**GCCA CCTCT CTT**CC** **C**ATTT TACTT G |
| **Alt.** | TCTGA TGTTT **CCCAC** **CC**AGC A**CCCC** **CC**AAC ACA**CC** **C**TGGG TT**CCC** **C**CCGC A**CCCC** TGCA**C** **CCCC**A GCCAG CCT**CC** **C**GCCA CCTCT CTT**CC** **C**ATTT TACTT G |

**SI Table 9**: Putative G4 sequences and variants for selected examples discussed in the main paper. The G-tracts or C-tracts in the G4 region are highlighted in bold, with 10 base pairs of flanking sequence shown on both sides.

| phenotype category | variant type | Total associated variants | Variants in linkage disequilibrium |
|---|---|---|---|
| *cis*-pQTL | common | 2634 | 1169 |
| *cis*-pQTL | rare | 212 | 4 |
| *trans*-pQTL | common | 14066 | 5781 |
| *trans*-pQTL | rare | 909 | 40 |
| binary | common | 1014 | 514 |
| binary | rare | 45 | 0 |
| quantitative | common | 8160 | 4804 |
| quantitative | rare | 325 | 10 |

**SI Table 10**: This table reports the number of non-coding variants in linkage disequilibrium ($R^2 \geq 0.2$) with nearby known GWAS variants (taken from NHGRI-EBI GWAS Catalog) located within a 1 Mb window (500 kb both sides of the variants) that are also associated with the same phenotype at genome-wide significance ($p < 10^{-8}$). Counts are stratified by variant frequency (rare, MAF $\leq 0.1\%$ and common) and by phenotype category. Complete results are provided in SI Table 11.

**SI Table 11**: This table reports the linkage disequilibrium ($R^2$) between all significantly associated non-coding variants and previously reported GWAS variants from the NHGRI-EBI GWAS Catalog located within a 1 Mb window (±500 kb of each variant). Only variant pairs in LD are reported. The table further includes a similarity score between the corresponding UK Biobank phenotype and the phenotype reported in the NHGRI-EBI GWAS Catalog, measured as Levenshtein distance (Methods).

| region | G4-disruptive | | G4-formative | | G4-modifying | |
|---|---|---|---|---|---|---|
| | **mean** | **median** | **mean** | **median** | **mean** | **median** |
| **All genes (with at least one G4-modifying variants)** | | | | | | |
| promoters (1 kb) | 10.8 | 8 | 5.2 | 3 | 67.1 | 40 |
| promoters (200 bp) | 6.6 | 5 | 3.3 | 2 | 39.6 | 25 |
| splice-proximal introns | 11.8 | 7 | 4.5 | 2 | 61.8 | 28 |
| UTR | 9.8 | 7 | 4.2 | 2 | 43.4 | 25 |
| UTRpro1Kbp | 15.9 | 11 | 7.4 | 4 | 97.8 | 59 |
| UTRpro200 | 12.2 | 9 | 5.6 | 3 | 71.0 | 43 |
| **Genes with significant associations at gene-level** | | | | | | |
| promoters (1 kb) | 14.3 | 13 | 13.7 | 7 | 116.9 | 62 |
| promoters (200 bp) | 9.2 | 10 | 21.5 | 7.5 | 45.7 | 37 |
| splice-proximal introns | 23.8 | 11 | 63.3 | 7.5 | 165.1 | 86 |
| UTR | 11.4 | 11 | 3.3 | 3.5 | 55.1 | 31 |
| UTRpro1Kbp | 20.5 | 15.5 | 12.3 | 7 | 128.4 | 72.5 |
| UTRpro200 | 16.4 | 14 | 13.3 | 5 | 72.5 | 46 |

**SI Table 12**: Summary of G4-disruptive, G4-formative, and G4-modifying variants across individual regulatory or combined regulatory regions corresponding to the gene-level collapsing models studied in this work (SI Table 13). The top portion reports the mean and median number of variants in regulatory regions containing at least one G4-modifying variant. The bottom portion presents the same statistics restricted to regulatory regions showing a significant association with any UK Biobank phenotype. For regions associated with multiple phenotypes, each region type is counted only once. Same information is plotted in SI Fig. 14.

| Category | Model | Qualifying Criteria |
|---|---|---|
| **Baseline non-coding models** | Rare variants | Collapses all rare variants within a specific gene regulatory region |
| | Rare variants CADD | Collapses rare variants with CADD score $\geq 5$ |
| **Mechanistic non-coding models** | G4-modifying rare variants | Rare variants that potentially modify G4 structures (i.e. includes all G4-disruptive, G4-formative, and G4-modulatory variants) |
| | G4-disruptive rare variants | Rare variants predicted to disrupt existing G4 structures |
| | G4-formative rare variants | Rare variants predicted to form new G4 structures |

**SI Table 13**: Five collapsing strategies were used in this study to aggregate rare non-coding variants. Each strategy was applied separately to promoters (1 kb), promoters (200 bp), UTRs, splice-proximal introns, UTR-promoters (1 kb), and UTR-promoters (200 bp), resulting in 30 collapsing models. In addition, for splice-proximal introns and UTRs, analogous template and non-template *G4-Mechanistic non-coding* models were constructed using G4 variants restricted to the template or non-template strands, yielding six additional models. In total, 36 collapsing models were evaluated based on five distinct collapsing strategies.

**SI Table 14**: Table (provided separately as an Excel file) listing the significant associations ($p < 10^{-8}$) identified in the gene-level collapsing PheWAS for protein abundances in the UKB dataset.

| **Collapsing models** | **Binary** | **Quantitative** | ***cis*-pQTL** | ***trans*-pQTL** |
|---|---|---|---|---|
| All rare variants | 0.76 (168) | 0.14 (154) | 0.40 (123) | 0.31 (324) |
| All rare variants (CADD≥5) | 1.02 (130) | 0.23 (170) | 0.65 (167) | 0.5 (284) |
| G4 rare variants | 1.43 (41) | 0.36 (73) | 0.86 (95) | 0.75 (287) |
| G4-disruptive rare variants | 0.0 (0) | 0.67 (46) | 0.96 (37) | 1.01 (116) |
| G4-formative rare variants | 2.93 (6) | 2.02 (8) | 0.71 (6) | 1.2 (67) |
| PTV | 1.34 (1435) | 0.66 (957) | 1.86 (691) | 1.19 (466) |

**SI Table 15**: Mean absolute effect sizes ($|\beta|_{\text{mean}}$) and (number of associations) for non-coding gene-collapsing models for significant ($p < 10^{-8}$) associations. All non-coding collapsing models are defined in SI Table 13. Data for PTV models are taken from refs. [1–3].

**SI Table 16**: Table (provided separately as an Excel file) listing the significant associations ($p < 10^{-8}$) identified from the gene-level collapsing PheWAS of binary traits in the UKB dataset.

**SI Table 17**: Table (provided separately as an Excel file) listing the significant associations ($p < 10^{-8}$) identified from the gene-level collapsing PheWAS of quantitative traits in the UKB dataset.

**SI Table 18**: Table (provided separately as an Excel file) listing the significant associations ($p < 10^{-8}$) identified from the gene-level (PTV + non-coding combined) collapsing PheWAS of protein abundances in the UKB dataset.

**SI Table 19**: Table (provided separately as an Excel file) listing the significant associations ($p < 10^{-8}$) identified from the gene-level collapsing (PTV + non-coding combined) PheWAS of binary traits in the UKB dataset.

**SI Table 20**: Table (provided separately as an Excel file) listing the significant associations ($p < 10^{-8}$) identified from the gene-level (PTV + non-coding combined) collapsing PheWAS of quantitative traits in the UKB dataset.

**SI Table 21**: Table (provided separately as an Excel file) listing AGD phenotypes considered in this study and their corresponding mappings to UKB phenotype codes. Where no mapping is provided, the phenotype is not available in the UK Biobank dataset.

**SI Table 22**: Table (provided separately as an Excel file) listing the significant associations ($p < 3 \times 10^{-9}$) identified from the gene-level collapsing PheWAS of binary traits in the AGD dataset (full cohort: 195,457 EUR samples).

**SI Table 23**: Table (provided separately as an Excel file) listing the significant associations ($p < 3 \times 10^{-9}$) identified from variant-level PheWAS of binary traits in the AGD dataset (full cohort: 195,457 EUR samples). This analysis has only been performed for phenotypes that are present in both AGD and UKB phenotypes.

**SI Table 24**: Table (provided separately as an Excel file) listing the significant associations ($p < 3 \times 10^{-9}$) identified from the gene-level collapsing PheWAS of binary traits in the AGD dataset (subset cohort: 117,016 EUR samples).

| | median | mean | sd | mean ± sd | mean ± 2sd |
|---|---|---|---|---|---|
| Variant-level (olink) | 1.014 | 1.023 | 0.047 | (0.976, 1.070) | (0.928, 1.118) |
| Variant-level (quantitative) | 1.033 | 1.044 | 0.141 | (0.903, 1.185) | (0.761, 1.327) |
| Variant-level (binary) | 1.002 | 1.003 | 0.052 | (0.951, 1.056) | (0.898, 1.108) |
| Gene-level (olink) | 1.005 | 1.006 | 0.040 | (0.966, 1.046) | (0.926, 1.086) |
| Gene-level (quantitative) | 1.016 | 1.019 | 0.063 | (0.957, 1.082) | (0.894, 1.145) |
| Gene-level (binary) | 0.995 | 0.992 | 0.118 | (0.874, 1.110) | (0.757, 1.227) |

**SI Table 25**: Genomic inflation ($\lambda$) statistics for variant-level and gene-level models across phenotype categories. Corresponding distribution plots are in SI Fig. 24.

| model | trait | significance cutoff | observed associations | permutated associations | FDR (%) |
|---|---|---|---|---|---|
| exWAS | olink | $p < 10^{-8}$ | 59926 | 13 | 0.022 |
| exWAS | binary | $p < 10^{-8}$ | 18267 | 15 | 0.082 |
| exWAS | quantitative | $p < 10^{-8}$ | 60228 | 3 | 0.005 |
| PheWAS | olink | $p < 10^{-8}$ | 1506 | 22 | 1.46 |
| PheWAS | binary | $p < 10^{-8}$ | 345 | 12 | 3.48 |
| PheWAS | quantitative | $p < 10^{-8}$ | 451 | 6 | 1.33 |

**SI Table 26**: Summary of false discovery rates (FDR) estimated using the $n$-of-1 permutation [1, 4] framework for variant-level and gene-level association analyses.

### Supplementary Figures

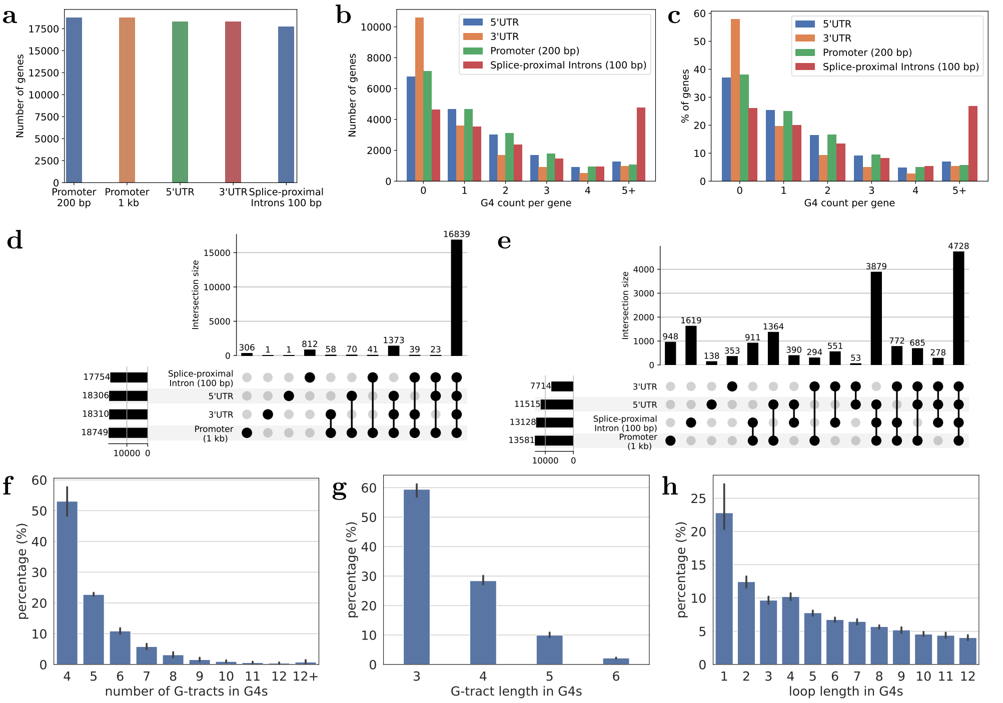


**SI Fig. 1**: Distribution of G4s and G4-modifying variants across non-coding regulatory regions. (a) Number of genes with regulatory regions (in GENCODE v43, autosomes only). (b–c) Distribution of G4s across genes. (d-e) UpSet plot showing the distribution of (d) genes with *regulatory regions* and (e) genes with G4s within their *regulatory regions*. In both UpSet plots, the left horizontal bars indicate the total number of genes with specific regulatory regions, whereas the top vertical bars indicate the size of each intersection, that is, the number of genes with multiple regulatory regions marked by connected black dots below. (f-h) Characteristics of G4 structures, including the number of G-tracts in G4s (f), and the lengths of G-tracts (g) and the lengths of loops (h) in *regulatory regions* of the human genome. Bars indicate standard deviation across *regulatory regions*.

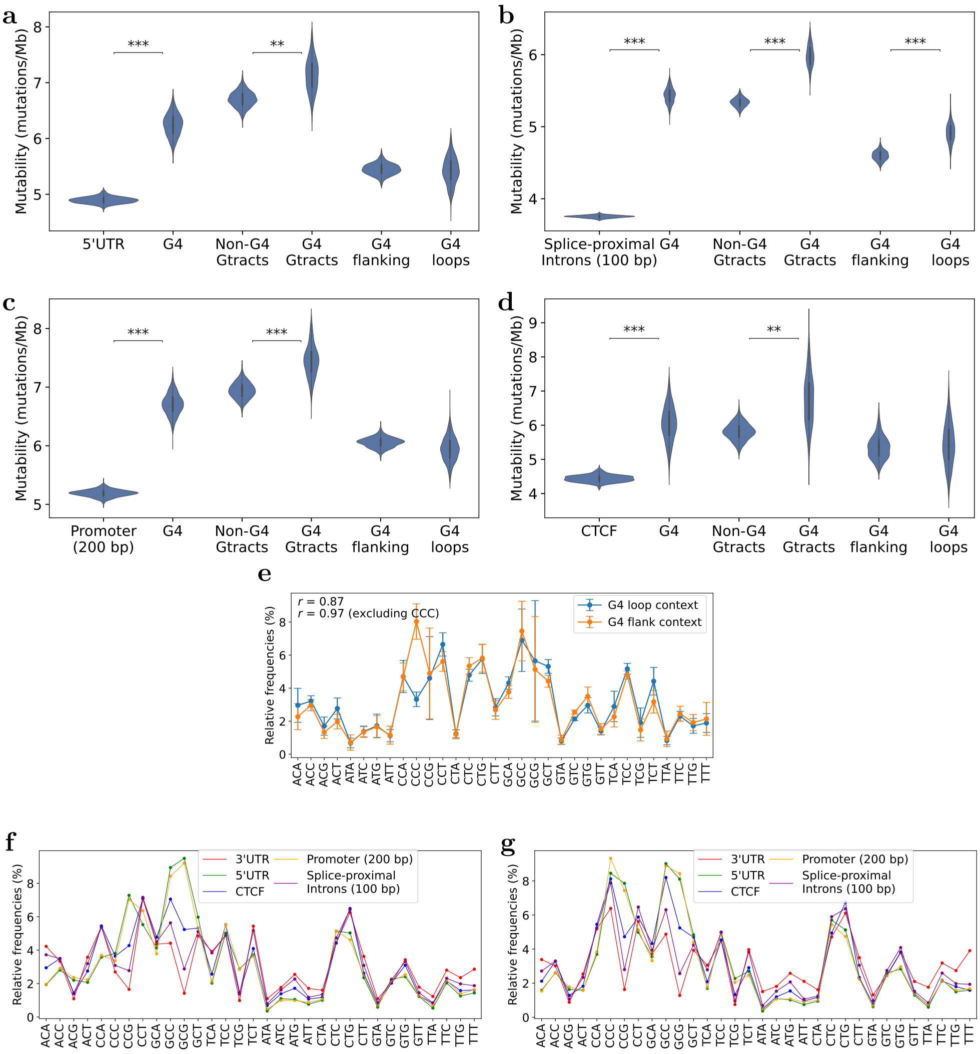


**SI Fig. 2**: Mutability (mutations/Mb) across non-coding regulatory regions including (a) 5′UTR, (b) splice-proximal introns, (c) promoters (200 bp), and (d) CTCF, computed considering only rare variants with minor allele frequency (MAF) $< 0.1\%$. The distribution is obtained through 1000 subsampling iterations and significance (two-sided t-test) is indicated by asterisks as follows: $*$: $p < 10^{-50}$, $**$: $p < 10^{-100}$, and $***$: $p < 10^{-150}$. (e) The plot illustrates the relative frequency of trimers in G4 loop and flanking sequences (100 bp on each side). Error bars show the standard deviation across *regulatory regions*. The Pearson correlation ($r$) between trimer frequencies in G4 loop and flanking sequences is 0.87, or 0.97 when excluding CCC. Trimer contexts in G4 (f) loops and (g) flanking sequence (100 bp on both sides).

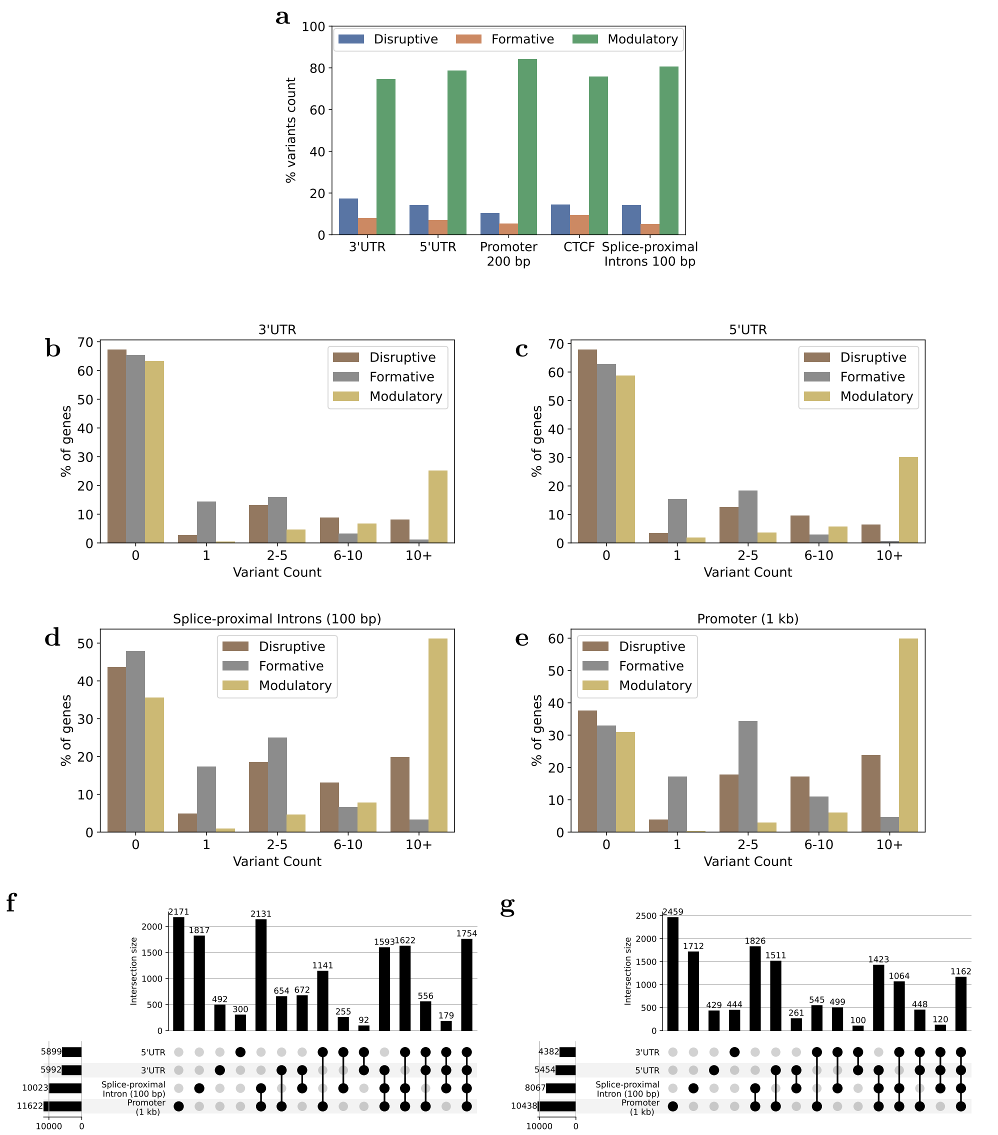


**SI Fig. 3**: (a) Proportion (%) of G4-modifying variants across *regulatory regions* with corresponding counts in SI Table 2. (b–e) Distribution of G4-modifying variants across non-coding regulatory regions including (b) 3′UTR, (c) 5′UTR, (d) splice-proximal introns, and (e) promoters (1 kb). UpSet plot showing the distribution of (f) genes with G4-disruptive variants within their *regulatory regions*, and (g) genes with G4-formative variants within their *regulatory regions*. In both UpSet plots, the left horizontal bars indicate the total number of genes with G4-disruptive (f) or G4-formative (g) variants in each regulatory region. The top vertical bars represent the size of each intersection, showing the number of genes with these variants across multiple regulatory regions, as indicated by the connected black dots below.

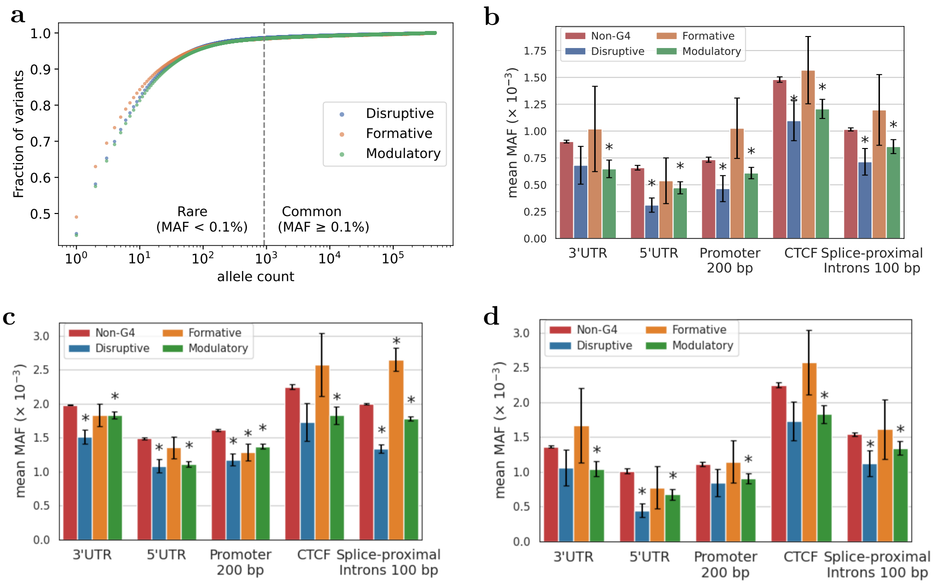


**SI Fig. 4**: (a) Allele frequency of G4-modifying variants. MAF of G4-modifying variants compared to non-G4 variants in the (b) UKB cohort for 1,389 intolerant genes, (c) Alliance for Genomic Discovery cohort, and (d) Alliance for Genomic Discovery cohort for 1,389 intolerant genes. Error bars denote the standard error of the mean. Intolerant genes were defined as protein-coding genes whose canonical transcripts had a loss-of-function observed/expected upper bound (LOEUF; *lof.oe_ci.upper*) $< 0.35$ based on constraint metrics from gnomAD v4.1. The $*$ indicates a statistically significant difference (Welch's t-test, $p < 0.05$) in mean MAF between G4-modifying and non-G4 variants within the same non-coding regulatory region.

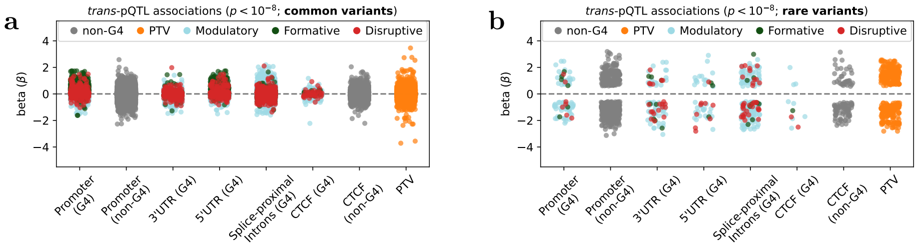


**SI Fig. 5**: (a-b) Effect size ($\beta$) distribution of *trans*-pQTLs for both common and rare (MAF $<$ 0.1%) variants in *regulatory regions*, alongside protein-truncating variant (PTV) associations [1–3]. Colored points denote different classes of variants (non-G4 non-coding, G4-modulatory non-coding, G4-disruptive non-coding, G4-formative non-coding, and protein-truncating coding variants).

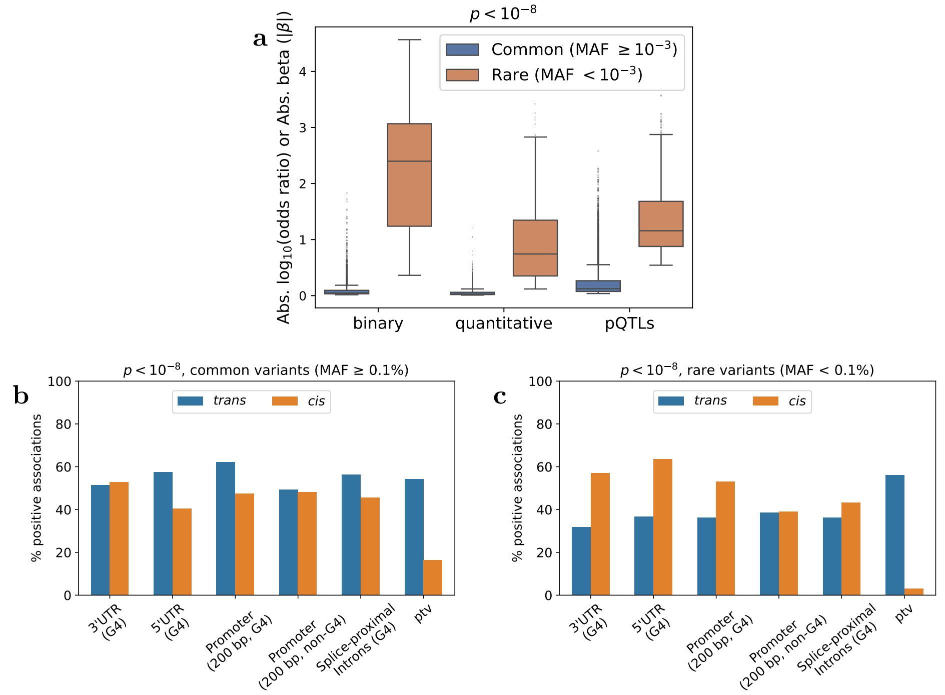


**SI Fig. 6**: (a) Box plot comparing the absolute effect size for significant ($p < 10^{-8}$) common versus rare non-coding variant associations. (b–c) Percentage of positive *cis*- and *trans*-pQTL associations at significance thresholds of $p < 10^{-8}$. Overall % positive association for PTV vs non-coding variants are summarised in SI Table 6.

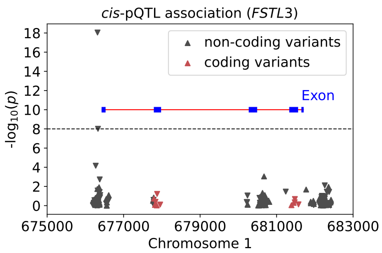


**SI Fig. 7**: Regional Manhattan plot for a representative cis-pQTL at *FSTL3* on chromosome 1. Points denote non-coding (grey) and coding (red) variants. The horizontal dashed line marks the genome-wide significance threshold. Triangle orientation denotes effect direction: downward triangles indicate decreased protein levels; upward triangles indicate increased protein levels.

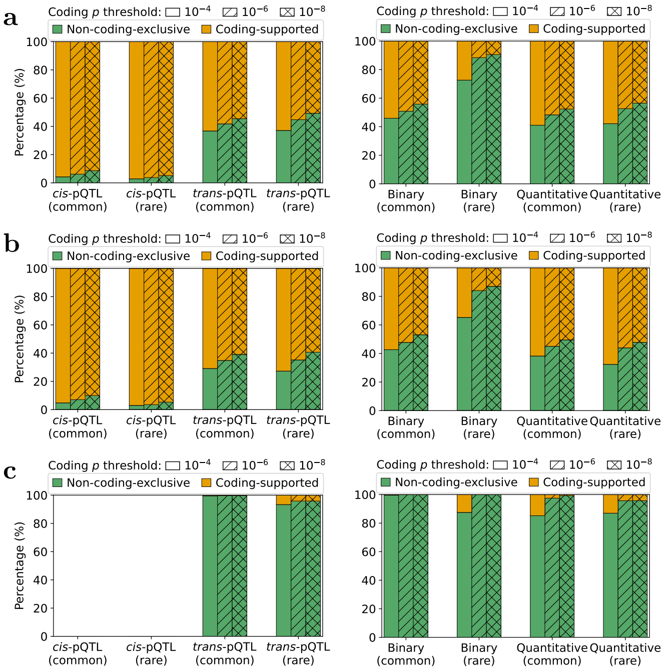


**SI Fig. 8**: Proportion of significant variant-level non-coding associations that are exclusive to non-coding models versus supported by coding variation (irrespective of effect direction) across *cis*-pQTL and *trans*-pQTL (left-panel) and binary and quantitative (right-panel) signals and stratified by allele frequency (common vs rare) for all non-coding variants in (a) *regulatory regions*, (b) promoter (200 bp) and (c) CTCF binding site. Bars are stacked by category: green indicates non-coding-exclusive associations; orange indicates coding-supported associations. Hatch patterns denote the coding significance thresholds ($p < 10^{-4}$, $p < 10^{-6}$, $p < 10^{-8}$).

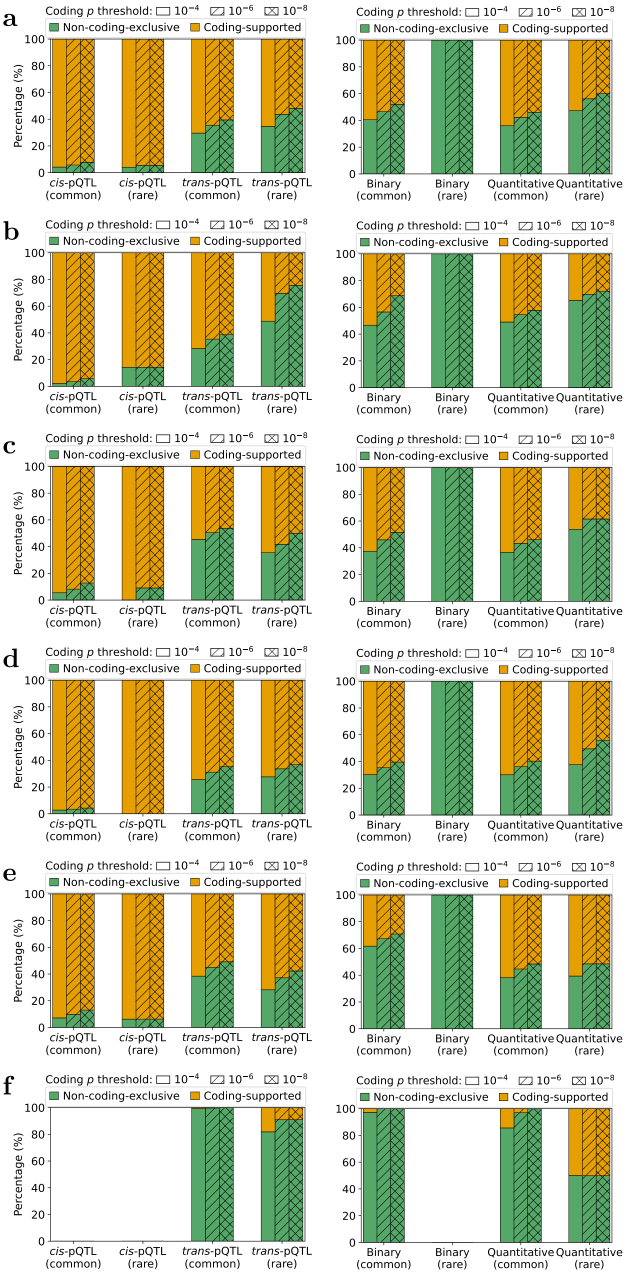


**SI Fig. 9**: Proportion of significant variant-level non-coding associations that are exclusive to non-coding models versus supported by coding variation (irrespective of effect direction) across *cis*-pQTL and *trans*-pQTL (left-panel) and binary and quantitative (right-panel) signals and stratified by allele frequency (common vs rare) for G4-modifying non-coding variants in (a) *regulatory regions*, (b) 3′UTRs, (c) 5′UTRs, (d) splice-proximal introns, (e) promoters (200 bp), and (f) CTCF binding sites. Bars are stacked by category: green indicates non-coding-exclusive associations; orange indicates coding-supported associations. Hatch patterns denote the coding significance thresholds ($p < 10^{-4}$, $p < 10^{-6}$, $p < 10^{-8}$).

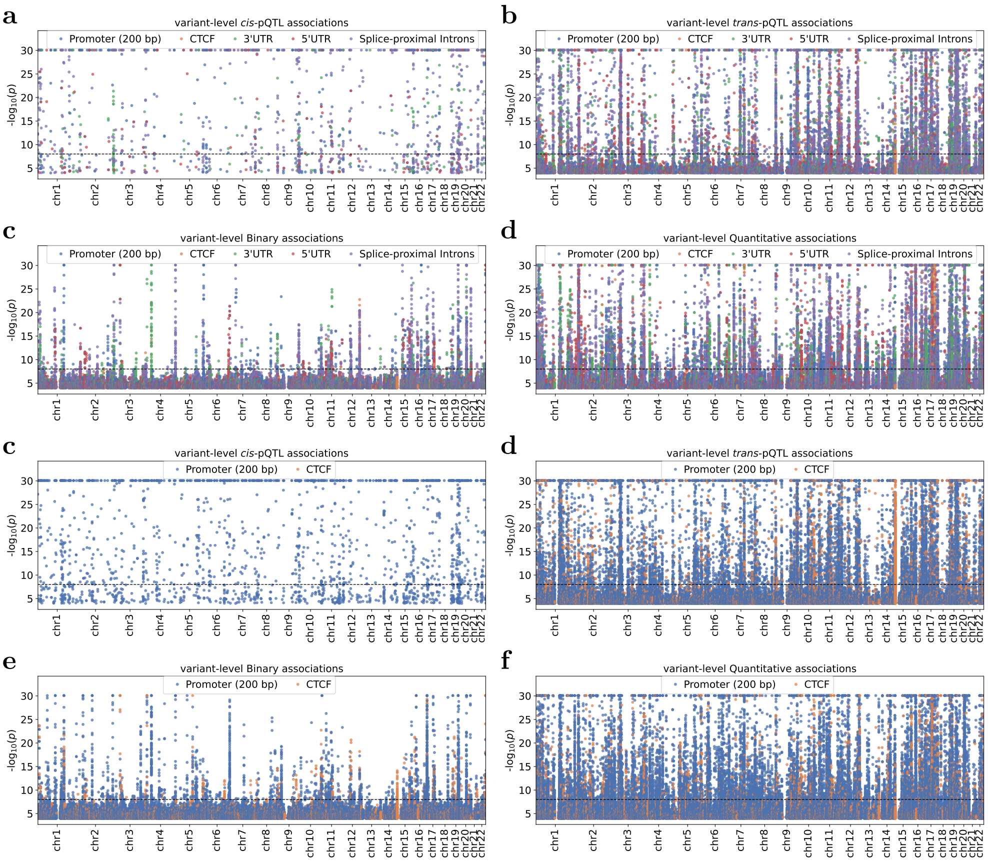


**SI Fig. 10**: Manhattan plots for variant-level associations of (a-d) G4-modifying variants with protein abundances (*cis* and *trans*), binary traits, and quantitative traits across *regulatory regions* and (e-h) non-G4 variants in promoters (200 bp) and CTCF binding sites with protein abundances (*cis* and *trans*), binary traits, and quantitative traits. The dashed line marks the study-wide significance threshold ($p < 10^{-8}$).

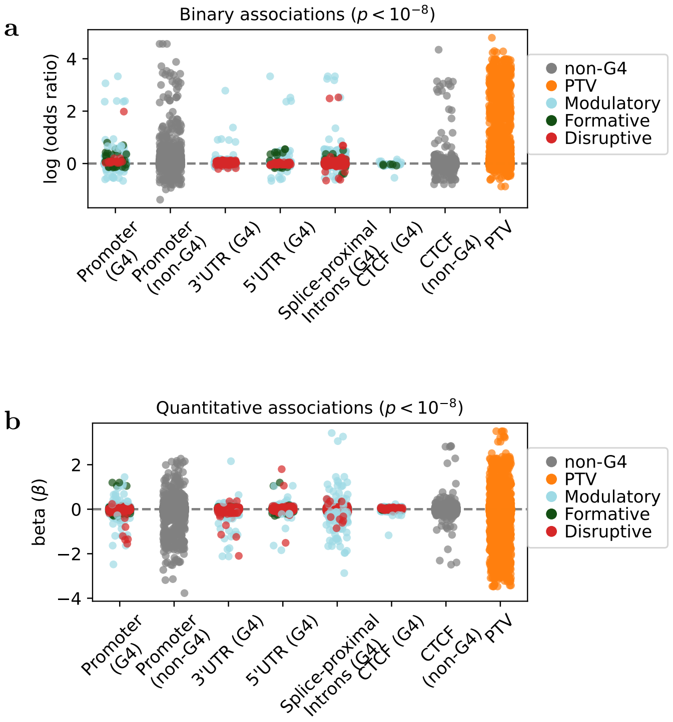


**SI Fig. 11**: Summary of variant–level non-coding associations. (a–b) Present effect sizes for significant binary (a) and quantitative (b) associations of variants in *regulatory regions*, alongside associations for protein-truncating coding variants. For promoters (200 bp) and CTCF binding sites, associations for non-G4 variants are also included.

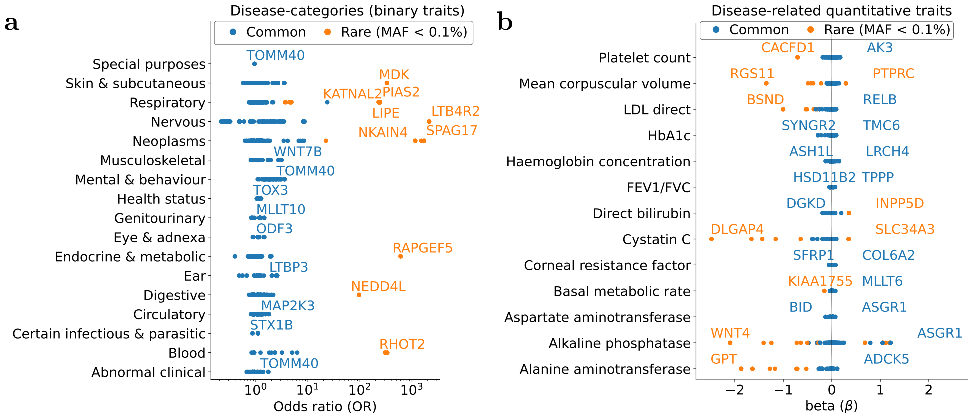


**SI Fig. 12**: Variant-level associations of non-coding variants across binary and quantitative traits. (a) Effect sizes for selected G4 variant binary associations per disease group ($p < 10^{-8}$), with labels for associations with the highest odds ratio in a disease group or odds ratio > 100. Abbreviations – FEV1/FVC: forced expiratory volume in 1 s/forced vital capacity ratio, HDL: high-density lipoprotein, and LDL: low-density lipoprotein. (b) Effect sizes of G4 variant associations for selected disease-related quantitative measures, with a dashed line at beta = 0. Rare and common variants are highlighted in orange and blue, respectively.

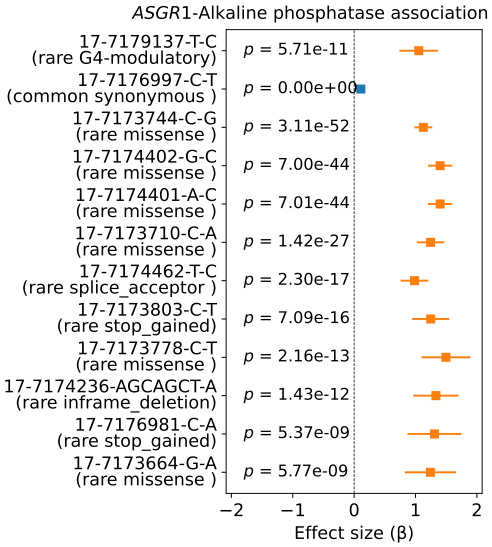


**SI Fig. 13**: Forest plots comparing effect size of coding and non-coding variants in *ASGR1* associated with alkaline phosphate. The G4 variant (17-7179137-T-C) is located in the 5′UTR (intronic) on the non-template strand of *ASGR1* and the complete G4 sequence is shown in SI Table 9. Rare and common variants are highlighted in orange and blue, respectively.

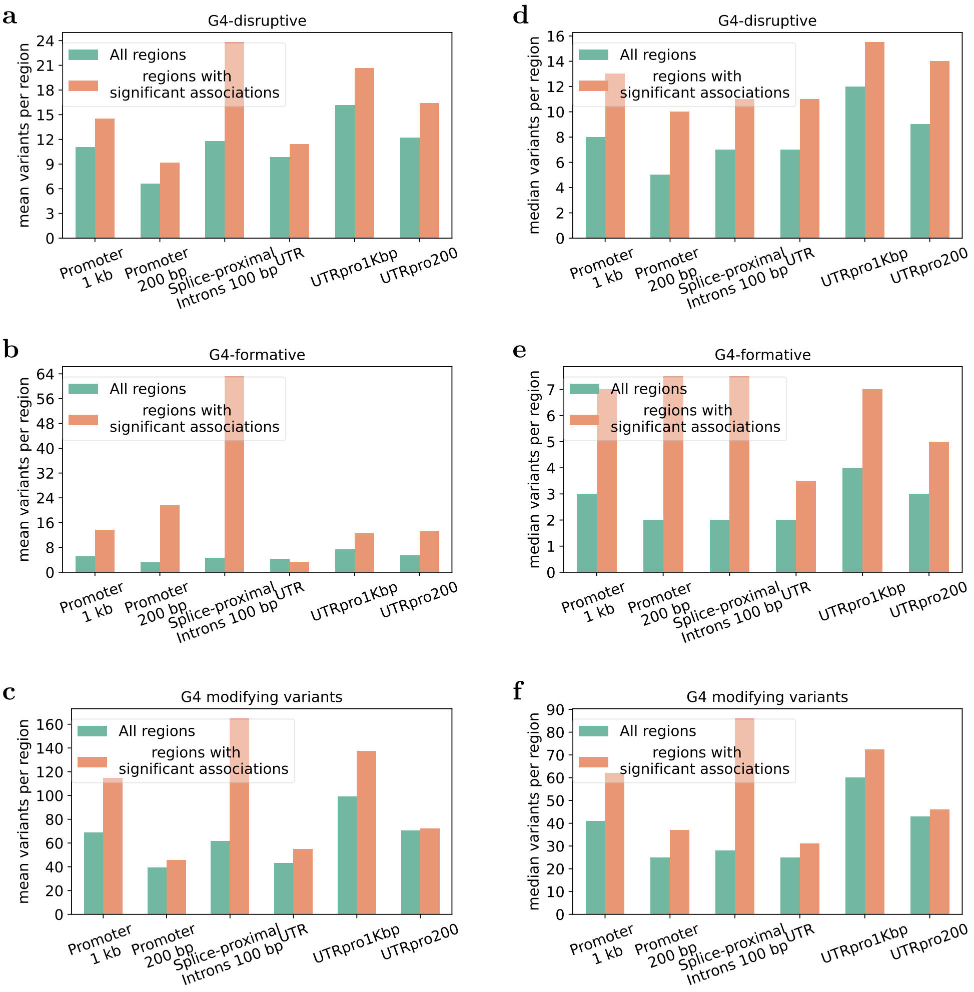


**SI Fig. 14**: Summary of G4-disruptive, G4-formative, and G4-modifying variants across individual *regulatory* or combined *regulatory* regions corresponding to the gene-level collapsing models (SI Table 13) studied in this work. The plot compares statistics for all regulatory regions (containing at least one G4-modifying variant, calculated per region) versus regulatory regions showing a significant association with any UK Biobank phenotype (SI Table 3). For regions associated with multiple phenotypes, each region type is counted only once. The same information is tabulated in SI Table 12.

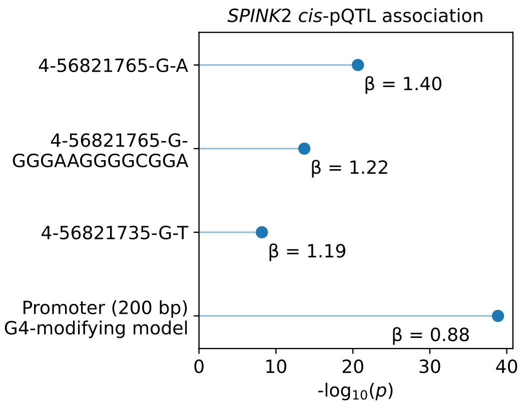


**SI Fig. 15**: Comparison of *SPINK2 cis*-pQTL signals in variant-level and gene-level association analyses for non-coding variants in promoter (200 bp). *G4-Mechanistic non-coding* collapsing models are detailed in SI Table 13.

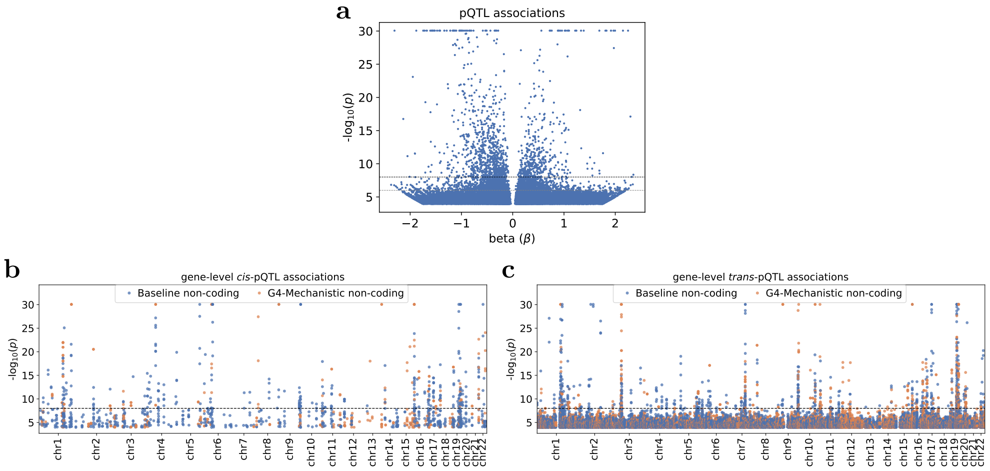


**SI Fig. 16**: (a) Volcano plot depicting all pQTL associations; association counts are summarized in SI Table 15. (b-c) Manhattan plots of gene-level collapsing associations for *cis*- and *trans*-pQTL associations. *G4-Mechanistic non-coding* and *Baseline non-coding* model associations are highlighted in two different colors. All the non-coding collapsing models are defined in SI Table 13.

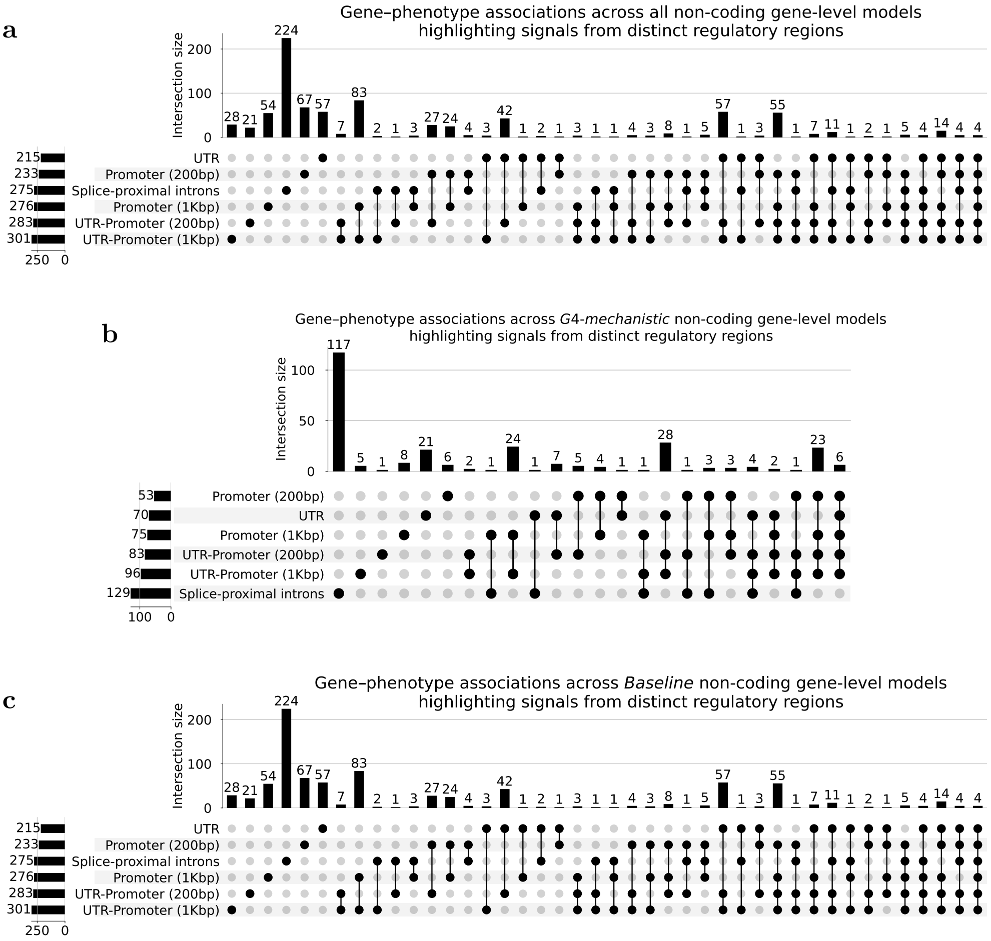


**SI Fig. 17**: UpSet plots demonstrating gene-phenotype associations across distinct regulatory regions for gene-level non-coding models: (a) all non-coding models, (b) *G4-Mechanistic non-coding* models, and (c) *Baseline non-coding* models. The plots highlight shared and region-specific association signals. In each UpSet plot, the left horizontal bars indicate the total number of associations detected in each individual regulatory region, whereas the top vertical bars indicate the size of each intersection, that is, the number of associations shared across combination of regions marked by connected black dots below. Definitions of all non-coding collapsing models are provided in SI Table 13. Total association counts include all phenotype categories considered in this study, including binary traits, quantitative traits, and protein abundances.

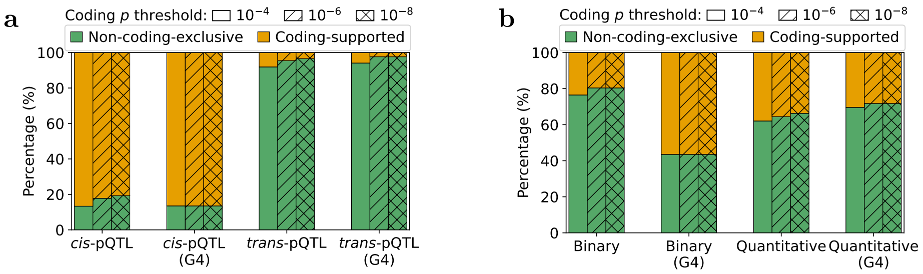


**SI Fig. 18**: Proportion of significant gene-level associations that are exclusive to non-coding models (SI Table 13) versus supported by coding-models across (a) *cis*-pQTL and *trans*-pQTL and (b) binary and quantitative signals and stratified by non-coding model type (all and *G4-Mechanistic non-coding*). Bars are stacked by category: green indicates non-coding-exclusive associations; orange indicates coding-supported associations. Hatch patterns denote the coding significance thresholds ($p < 10^{-4}$, $p < 10^{-6}$, $p < 10^{-8}$).

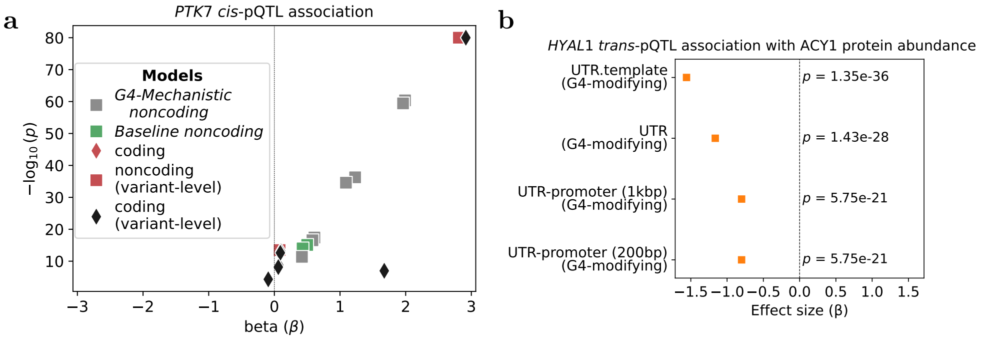


**SI Fig. 19**: (a) Comparison of *PTK7 cis*-pQTL signals in variant-level and gene-level association analyses for both coding and non-coding variants. (b) Forest plot comparing effect sizes across multiple coding and non-coding models for *trans*-pQTL associations of *HYAL1* with ACY1 protein abundance. *Baseline* and *G4-Mechanistic non-coding* collapsing models are detailed in SI Table 13 and coding-models [3] are briefed in the Methods.

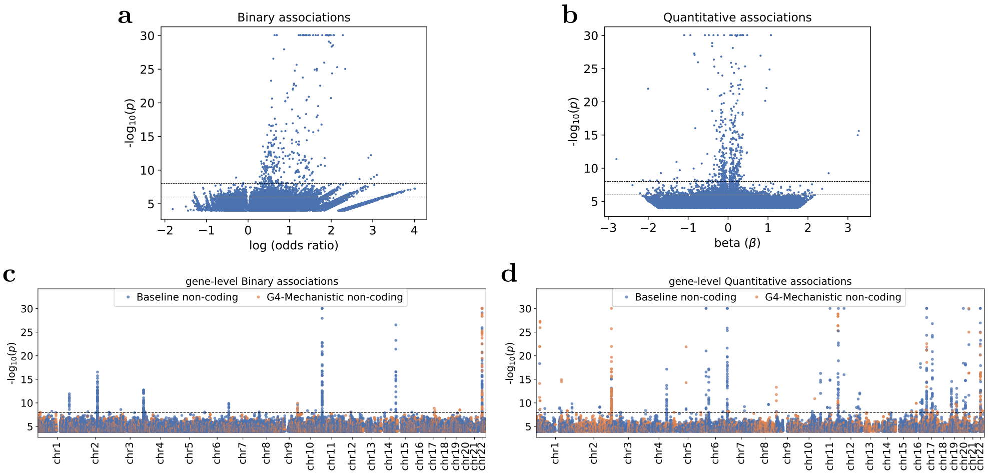


**SI Fig. 20**: (a-b) Volcano plot depicting all binary and quantitative associations; association counts are summarized in SI Table 15. (c-d) Manhattan plots of gene-level collapsing associations for binary and quantitative associations. *G4-Mechanistic non-coding* and *Baseline non-coding* model associations are highlighted in two different colors. All the non-coding collapsing models are defined in SI Table 13.

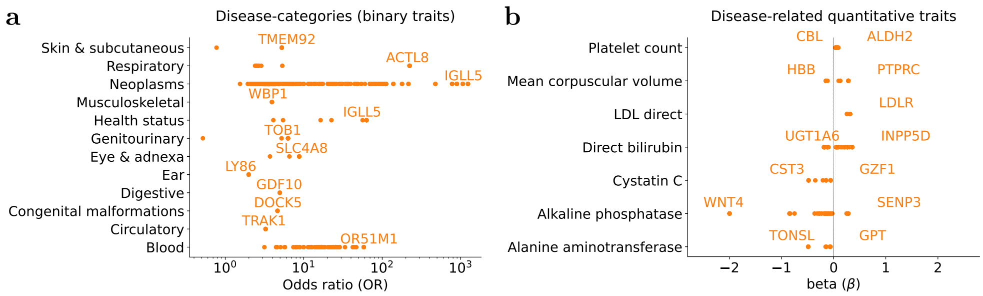


**SI Fig. 21**: Non-coding gene-level collapsing model association summary. (a) Effect sizes for key gene-disease associations ($p < 10^{-8}$), with labels for associations with the highest odds ratio in a disease group or odds ratio > 100. (b) Visualization of major gene–trait associations for selected disease-related quantitative measures. Abbreviations – LDL: low-density lipoprotein.

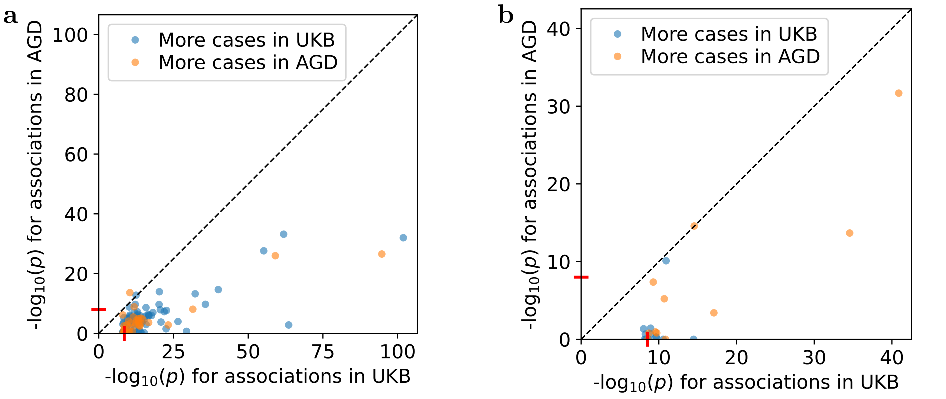


**SI Fig. 22**: (a) Concordance of $p$-values for gene-level associations observed in both cohorts. 121 of 123 significant UKB associations are shown (two AGD tests excluded due to $< 5$ cases/carriers). The significance threshold for AGD associations was set to $3 \times 10^{-9}$ (FDR $= 3.03\%$) to align with the UKB gene-level binary threshold (FDR $= 3.48\%$). The significance threshold is indicated by red ticks on the x- and y-axes. (b) Concordance of $p$-values for shared promoter variant-level associations between these two cohorts. This analysis was restricted to the 4,367 binary phenotypes shared between the UKB and AGD cohorts, and included only genotypes available in both cohorts.

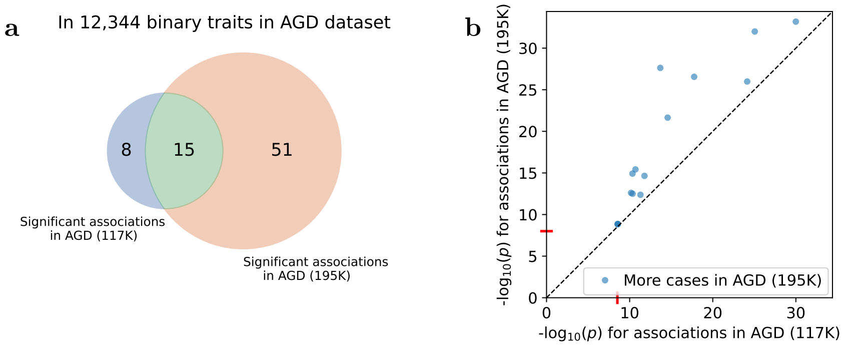


**SI Fig. 23**: (a) Venn diagram showing the overlap of significant gene-level associations in *regulatory regions* across 12,344 binary traits in AGD 117K (subset cohort) and AGD 195K (full cohort). (b) Comparison of $p$-values for shared significant associations between these two sets. The significance threshold is indicated by red ticks on the x- and y-axes.

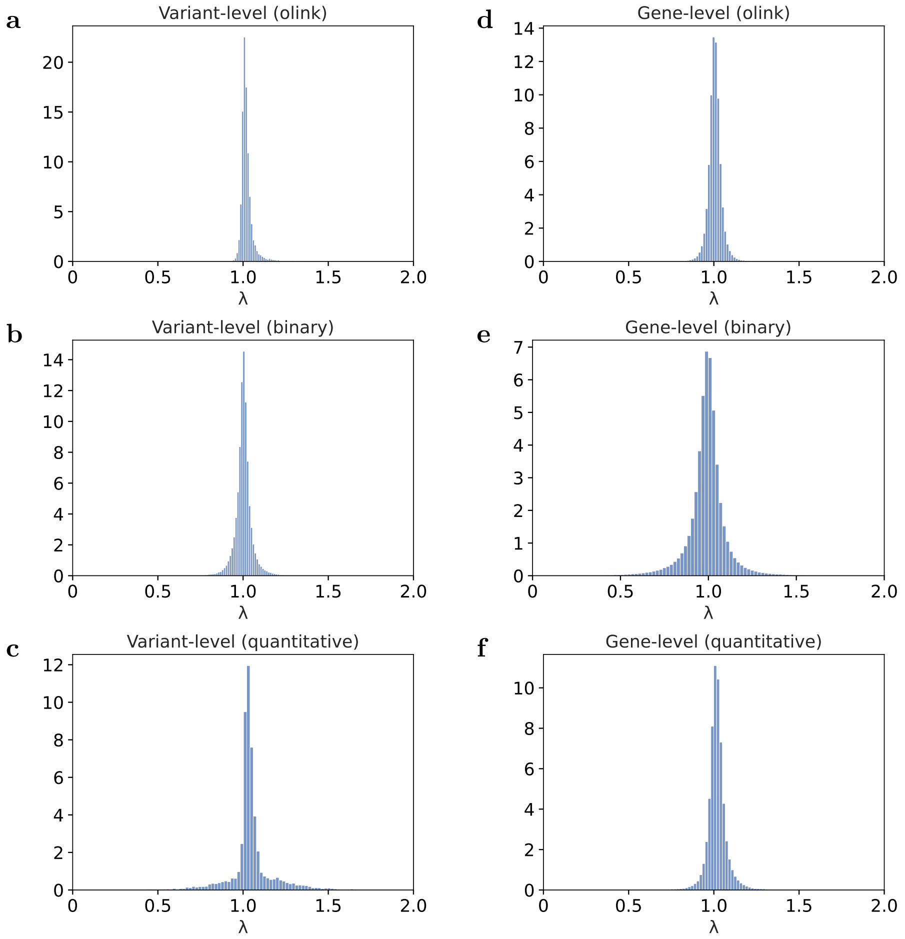


**SI Fig. 24**: Distribution of genomic inflation ($\lambda$) for variant-level and gene-level collapsing models across phenotype categories. Corresponding summary statistics are in SI Table 25.
